# Scarring and Selection in the Great Irish Famine

**DOI:** 10.1101/2020.09.07.20189662

**Authors:** Matthias Blum, Christopher L. Colvin, Eoin McLaughlin

## Abstract

What is the health impact of famines on survivors? We use a population exposed to severe famine conditions during infancy to document two opposing effects. The first: exposure leads to poor health into adulthood, a scarring effect. The second: survivors do not themselves suffer health consequences, a selection effect. Anthropometric evidence on over 21,000 subjects born before, during and after the Great Irish Famine (1845-52), among modern history’s most severe famines, suggests selection is strongest where mortality is highest. Individuals born in heavily-affected areas experienced no measurable stunted growth; scarring was found only where excess mortality was low.

## 1. Introduction

One of the universal features of famines is the immediate and devastating human tragedy they bring. Another, apparently, is the subsequent deterioration in the health and human capital of survivors. A plethora of case studies already document the scale of catastrophic famine events measured using adult stature. Figure 1 depicts our review of these other anthropometric studies. We plot famine-period excess mortality against the impact on the average societal stature of the famine-born cohort. The figure reveals the smallest drops in stature occur in populations exposed to the most severe incidents of famine-induced hunger and disease. This highlights the fact that surviving populations are selected populations which have fundamentally different characteristics to those that died or moved away, something that the studies depicted in the figure do not themselves tend to dwell on very much. But it is the health and human capital of such selected residual populations that is crucial in understanding a society’s subsequent recovery. Those that perished, or migrated away, do not, by definition, contribute directly to the wealth of the nation. Our contribution is our ability to use individual-level data to investigate this selection effect.

**Figure 1:**
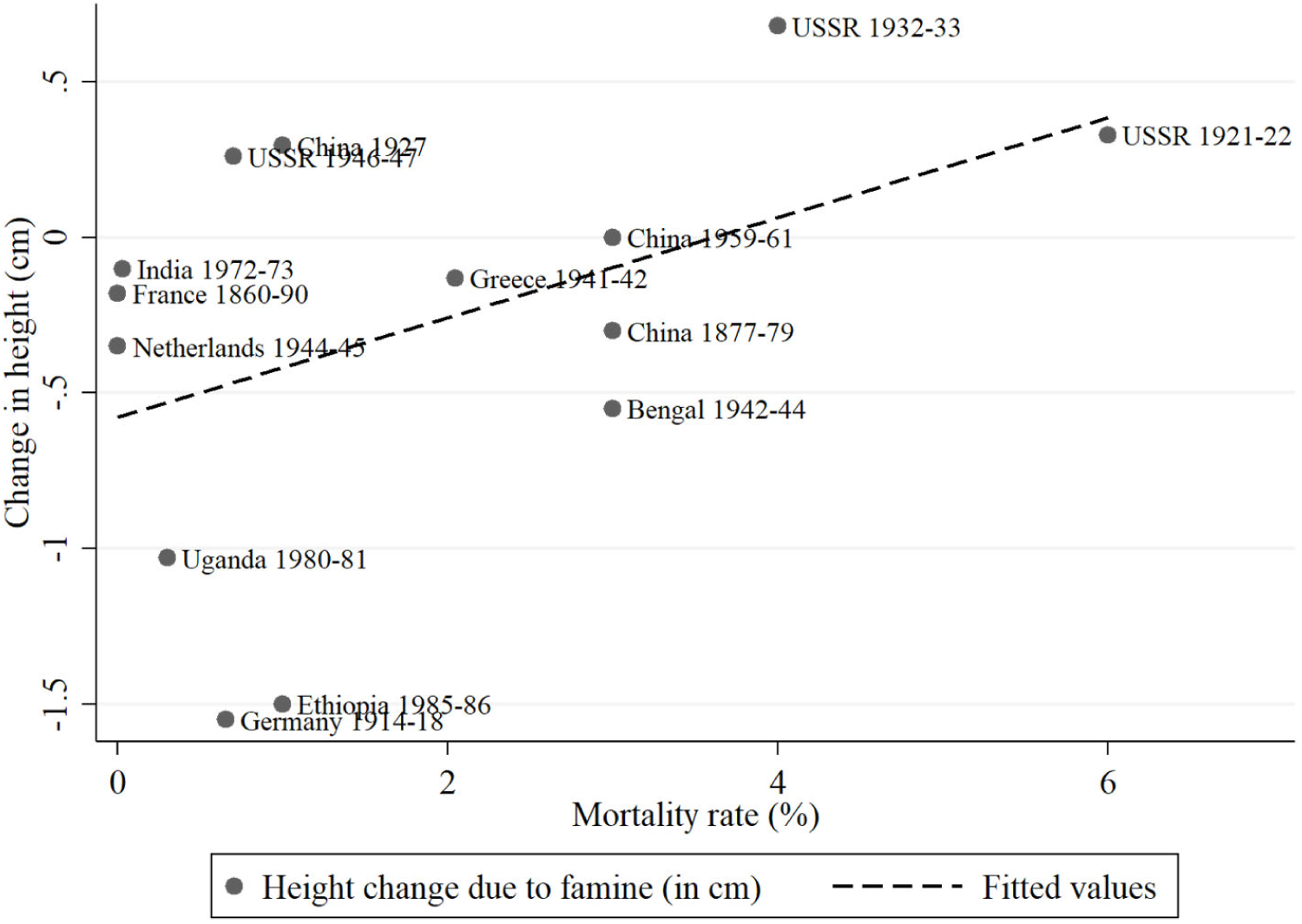
Change in average adult height and mortality in 14 famines *Sources:* Excess mortality figures: Ó Gráda (2007); corresponding height changes, defined as the difference in final adult height between pre-famine and famine-born cohorts: Bengal 1942-44 (Guntupalli and Baten 2006); China 1877-79 (Baten et al. 2010); China 1927 (Morgan 2004); China 1959-61 (Gørgens et al. 2012); Ethiopia 1985-86 (Dercon and Porter 2014); France 1860-90 (Banerjee et al. 2010); Germany 1914-18 (Blum 2011); Greece 1941-42 (Valaoras 1970); India 1972-73 (Guntupalli 2007); the Netherlands 1944-45 (Van Wieringen 1972); Uganda 1980-81 (Umana-Aponte 2011); USSR 1921-22, USSR 1932-33 and USSR 1946-47 (Wheatcroft 2009).

We study one of the most severe and lengthy famines in modern history, The Great Irish Famine of 1845-52: one million people died, and another million emigrated (Mokyr 1985). When deaths are considered relative to overall population levels, the Great Irish Famine overshadows practically all other famines: 12 per cent of Ireland’s population perished (Ó Gráda 2007: Tab. 3). This famine also led to devastating structural change: the catastrophe meant that the island’s potato-based economy was simply no longer viable, and it intensified pre-existing patterns of migration from which the island has not yet recovered. We assess the long-run consequence of this catastrophic event on the health and wellbeing of those born in famine conditions, remained in Ireland and survived into adulthood.

Adult stature reflects a population’s living standards in early life, with the crucial period being from inception to the age of two (Currie and Vogl 2013).^1^ The average adult height of the generation born in distressed conditions is a measure of a society’s health capital used widely by health and development economists – and economic historians – researching catastrophic events (Fogel 2004; Floud et al. 2011; Deaton 2013). The biological mechanisms underpinning the link between early childhood exposure to acute malnutrition and reduced adult stature relate to fetus growth *in utero*.^2^ Known as the “Barker thesis”, or “fetal origins hypothesis” (FOH), scientists link low birth weight with a variety of poor health outcomes into adulthood besides short stature (Barker 1990, 2004; Calkins and Devaskar 2011; Almond and Currie 2011). Scholars use adult stature as a measurable proxy for these other health conditions.

Continuing in this anthropometric tradition, we focus on average societal stature as our measure of health and human capital in the context of the Great Irish Famine (henceforth referred to simply as “the Famine”). Our individual-level data contain over 21,000 subjects born before, during and after the Famine in locations across Ireland which varied in their exposure to famine-induced nutritional deprivation and disease. These micro-data are drawn from archival records of prisons located in Dublin, Ireland’s capital city, and Clonmel, in rural Tipperary. Indeed, one of our contributions is the collection and use of these fine-grained data; previous explorations of this, and other, famines do not enjoy such large individual-level datasets.

The Famine’s root cause was an exogenous ecological shock – *Phytophthorainfestans* (potato blight) – which decimated the main source of food of the majority of the population. Those born during the Famine suffered this ecological shock in a crucial developmental stage and experienced devastating and often permanent health consequences, ranging from chronic disease to early death. The catastrophic event led to structural changes in food production as the potato was rendered an unreliable source of sustenance until the end of the nineteenth century, when the (Bordeaux mixture) treatment for potato blight was first discovered and applied. Consequently, those born after this famine enjoyed a very different diet than earlier generations.

Famines exacerbate the mortality of specific elements of the population (young and old). They also cause reductions in birth rates as people are unwilling or unable to procreate. Afterwards, birth rates rebound (Ó Gráda 2009). Both the selection in mortality and fertility are issues which can affect the attributes of a population after a famine. A poor childhood nutritional and disease environment can therefore have two effects that work in opposite directions: scarring and selection (Deaton 2007; Bozzoli et al. 2009). In the scarring outcome, surviving children do not realise their adult height potential, with the degree of the reduction depending on the severity of the nutritional and disease shock. This, in turn, may lower societal average height. By contrast, selection occurs when a high disease and low nutritional environment raises the survival cut-off threshold and fewer (low-height/low-health) children survive into adulthood, potentially *increasing* the average health and height of survivors (Alter 2004).

Our principal contribution derives from our historical context, which is uniquely suited to the study of catastrophic events in general, and famines in particular, because the strong spatial heterogeneity in its impact permits us to conduct an ecological identification of exposure during early childhood. Our micro-study offers new insights because competing scarring and selection effects may be hidden in the aggregated national studies typical of this literature. We contribute by attempting to differentiate scarring from selection effects. Indeed, we find the same famine severity/health capital correlation depicted in Figure 1 occurring within this one famine episode. Most famine deaths were recorded in rural parts of Ireland; direct famine-related mortality in Dublin, the island’s largest urban centre, was absent. And while we find few signs of early childhood malnutrition in the surviving population born in rural Tipperary, we *do* find evidence of such scarring of those born in Dublin (Figure 2). Our estimates of the extent of this scarring range between 0.3 cm (Table 4, Model 1) and 2.5 cm (Table C1, Model 1).^3^ These estimates represent large biological and economic effects: Baten and Komlos (1998) associate a 1 cm decline with a reduction in average life expectancy of 1.8 years; McGovern et al. (2017) link a 1 cm decrease with a 4 per cent fall in wages.

**Figure 2:**
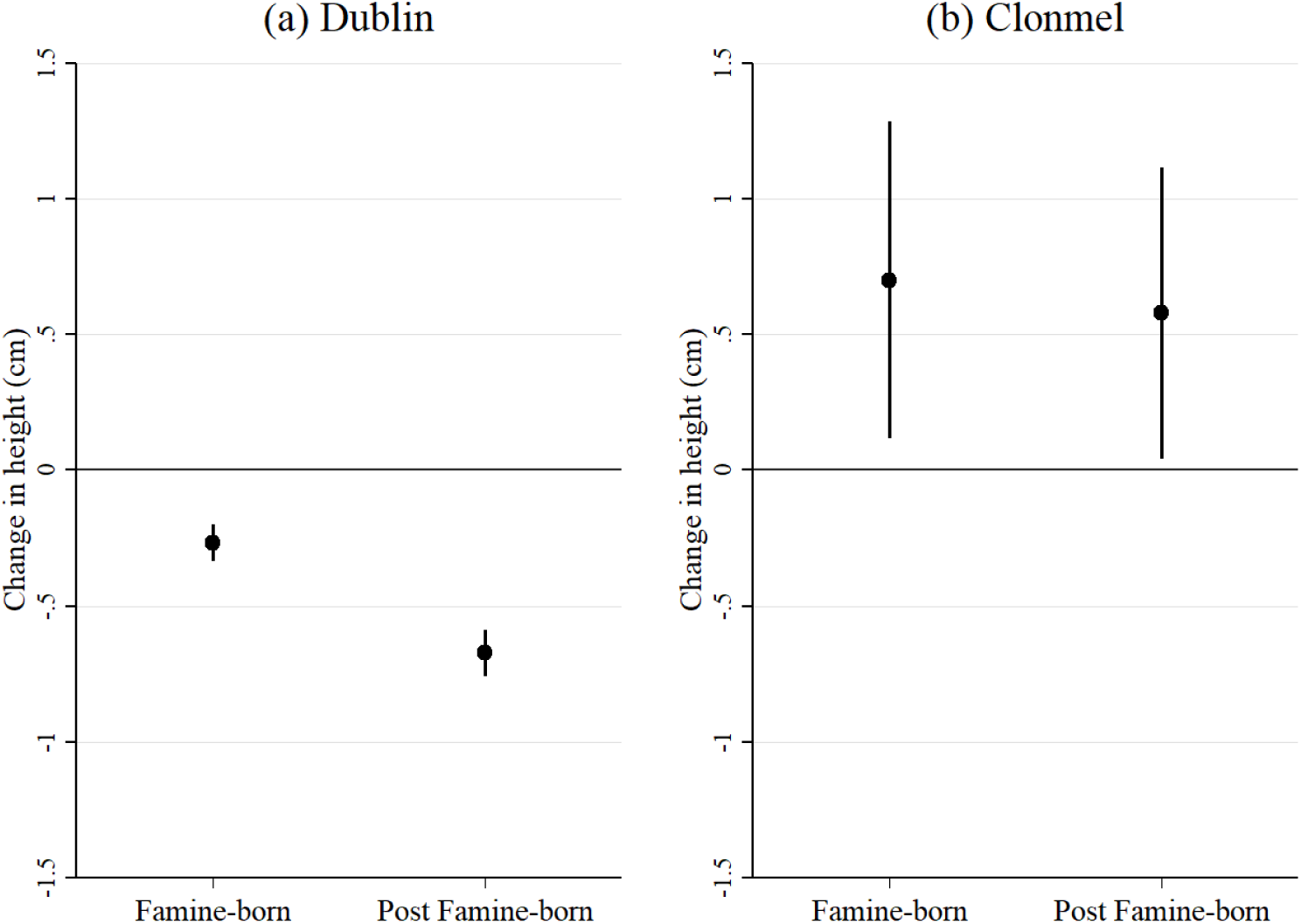
Change in average adult height in the Great Irish Famine *Notes:* Error bars depict 95% confidence intervals; reference category is an adult, pre-famine-born illiterate male Catholic who committed an offence against property (non-violent) and was imprisoned before 1877; see Table 4.

Famines are not just a historical curiosity; policymakers are once again interested in understanding the impact of this catastrophic risk. Historically, absolute food availability was more important than distribution and entitlement issues in causing famines (Alfani and Ó Gráda 2018; cf. Sen 1977). Extreme climate change may mean the same becomes the case in future. We contribute by showing how a historical case can be used to think about questions of potentially contemporary relevance.

This paper proceeds as follows. Section 2 provides the necessary background on the Great Irish Famine. Section 3 sets out the issue of selection in individual-level data and how this affects studies of the FOH. Section 4 outlines our data source. Section 5 defines our empirical strategies, while Section 6 presents our main findings, and Section 7 introduces a set of additional tests. Section 8 discusses our results and restates our overall contribution. A set of appendices report on the details of a series of robustness exercises and extensions.

## 2. Ireland’s Famine

Ireland experienced Western Europe’s last great subsistence crisis: back-to-back harvest failures between 1845 and 1852 caused by the fungus *Phytophthora infestans*, or potato blight. Many farmers had come to rely exclusively on the Irish Lumper variety of potato because it was particularly well suited to the island’s poor-quality soil (Bourke 1993). The Lumper proved to be very susceptible to blight, and there existed few alternative sources of sustenance for the majority of rural folk; rural Ireland had become a monoculture. Approximately 20 per cent of Ireland’s population either perished or migrated as a result of this blight-induced famine, reducing the island’s population from 8.17 to 6.53 million between the recorded censuses of 1841 and 1851 (BPP 1851). Then, in the post-Famine period, the potato became a much less reliable source of food security due to the continued presence of blight. As a consequence, Ireland’s diet shifted to wheat-based sources of carbohydrates (Clarkson and Crawford 2001).

Mokyr and Ó Gráda (2002) estimate about half of the deaths during the Famine were associated directly with malnutrition, while the other half were caused by indirect effects on personal behaviour and social structure. There were principally five famine-related illnesses: dysentery, diarrhoea, dropsy, starvation, and “fever” - typhoid, typhus and relapsing fever (MacArthur 1956: pp. 265-68). Famine mortality figures were highest in Munster and Connacht (west and south-west of the island), followed by Leinster (east). The lowest mortality was observed in Ulster (north-east). Figure 3 locates the regional impact of the Famine in terms of excess mortality, using county-level estimates from Mokyr (1985).

**Figure 3:**
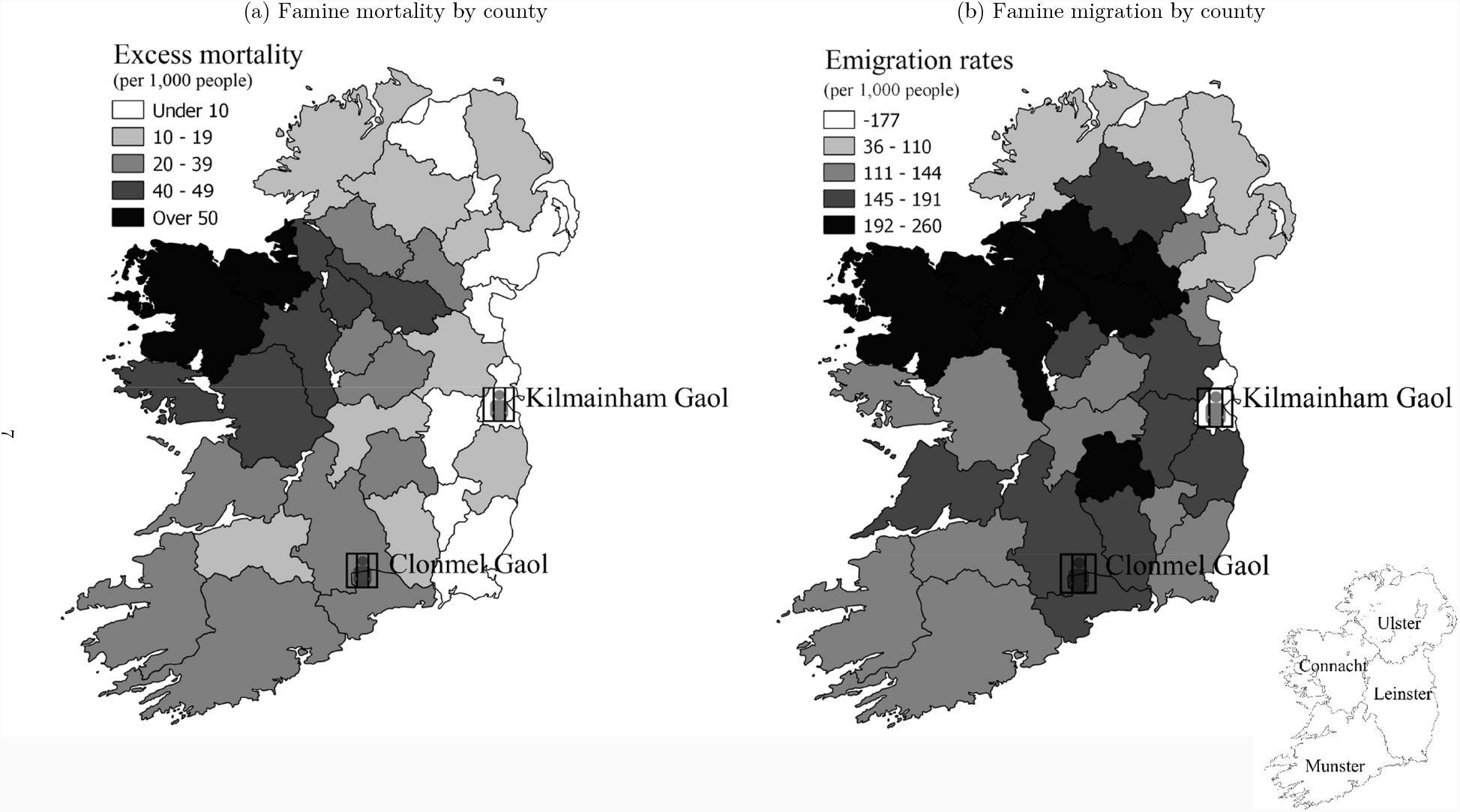
Famine maps of Ireland, with geographic location of two prisons *Notes:* Excess mortality rates are an average annual estimated total; emigration rates are totals expressed relative to the 1841 population. *Sources:* Mortality data from Mokyr (1985), migration data from Ó Gráda and O’Rourke (1997), and GIS shapefile adapted from Gregory and Ell (2004).

In terms of this study, the relevant populations relate to the counties in which our subjects were recorded, and earlier, born. We have individuals born across the island. But the majority hail from two locations: the Munster county of Tipperary, and the Leinster county of Dublin. Contrasting population trends can be seen. The population of Tipperary was ravaged by the Famine: in 1841 the population stood at 0.44 million, but by 1851 this had fallen by a quarter to 0.33 million. Lower and upper bound estimates of excess mortality in Tipperary during the Famine are 24 and 35 per 1,000 population (Mokyr 1985). This compares to the all-island median of between 19 and 26 deaths per 1,000 (see Table 1).

**Table 1:** Key demographic indicators, 1841-1851

|  | Dublin | Tipperary | Ireland |
| --- | --- | --- | --- |
| $\Delta$ population % (1841-1851) | 8.68 | -23.87 | -19.96 |
| Excess mortality per 1,000 pop. (lower bound) | -2.1 | 23.8 | 24.3 |
| Excess mortality per 1,000 pop. (upper bound) | 0.7 | 35.0 | 33.0 |
| $\Delta$ births per 1,000 population (1841-1851) | -1.65 | -6.93 | -4.44 |
| Cohort depletion % (1841-1851) | 13.12 | -31.21 | -30.82 |

*Notes and Sources:* Excess mortality estimates taken from Mokyr (1985: Tab. 9.1 and 9.2). Births per 1,000 population taken from census returns of those under the age of one at time of census and do not include adjustments for infant mortality. Cohort depletion refers to the percentage ratio of the cohort aged 5-14 from the 1841 census, relative to the 15-24 cohort in 1851. This measure includes the combined effects of changes to mortality, fecundity and migration.

Meanwhile, Greater Dublin (city and surrounding county) had a population of 0.37 million in 1841. This increased slightly to 0.40 million by 1851 as migrants from elsewhere in Ireland fled the famine conditions. The increase in the population of Greater Dublin masks the growing “slumification” of the city. Dublin’s plight was somewhat atypical for cities in the UK in the

Nineteenth century as its population was not sustained by growing industrialisation; the city was increasing in population size without consummate improvement in the quantity or quality of its housing stock. The slumification also had an influence on the general disease environment, so much so that Dublin had one of the lowest life expectancies of comparable cities (Prunty 1998).

As the population of Greater Dublin expanded over the decade 1841-51, an older strand of literature saw Dublin (as well as Cork City and Belfast) as being ‘unaffected directly’ by the Famine (Lynch and Vaizey 1960). Ó Gráda (1997: pp. 157-193) challenges this view, but concedes the famine mechanism was indirect. One such mechanism was increases in food prices and resulting falls in entitlements; the inhabitants of Dublin were less likely to be affected by an absolute decline in food availability as the urban diet was as much based on wheat (bread) as potatoes (Ó Gráda 1999: p. 163). Data on urban wages suggest a slight decline in real terms (D’Arcy 1989; Kennedy and Solar 2007). The second mechanism was immigration of famished rural-folk into the city. Indeed, Ó Gráda (1999) finds any increased urban mortality during the peak famine years was not driven by native Dubliners; rather, it was caused by rural immigrants dying of fever and dysentery. Together, these indirect famine effects make the Dublin-born population a useful comparison group for the directly-exposed rural population of Tipperary.

Internal and international migration was one of the main non-governmental responses to famine conditions (Kenny 2000; Donnelly 2001; Ó Murchadha 2011). Migration enabled the famished to go to sources of food located in urban centres in Ireland and beyond. The Irish-born population of Britain almost doubled between 1841 and 1851, from 415,725 to 727,326 (Macraild 2006). North America was the main destination for international migration (Akenson 1996). After the Famine, Ireland had the highest migration rates of Western Europe (Hatton and Williamson 1993, 1994). As with famine-related mortality, famine-induced migration differed regionally. Ó Gráda and O’Rourke (1997) estimate Tipperary experienced one of the highest emigration rates relative to mortality, with a ratio of 1.18, whereas Dublin was an anomaly in the sense it was a net-recipient of migrants (see Figure 3).

## 3. Understanding Selection

In Lumey et al.’s (2011) comprehensive review of the effects of famine on prenatal and adult health outcomes, the most consistent findings across studies relate to body size (height and BMI), diabetes and schizophrenia. The reviewed studies share many similarities with our own: they are observational and not experimental in nature; they focus on outcomes at future points in time; they are cohort studies; and they use population-based registries to trace people. Lumey et al. (2011) outline the analytical strategies which have been applied to date, including simple cohort analysis, difference-in-difference estimation and sibling designs. Across studies, definitions of famine exposure are ecological rather than based on individual food consumption as such data are unavailable. Primary exposure to food shortages are difficult to disentangle from other associated famine effects within any one famine episode. Furthermore, chronologies of famines vary from the well-defined (the Dutch Hongerwinter famine lasted exactly six months) to the much-less-so (seasonal famines in The Gambia between 1949 and 1994).

A key issue in the literature on the health effects of early-life exposure to famine relates to scarring and selection, two forces which can co-occur, but to different degrees depending on the severity of the famine. In the literature thus far, the issues surrounding competing scarring and selection effects has probably been most evident in studies of the Leningrad siege. In their analysis of a sample of survivors of this 872-day German siege of St Petersburg during WWII, Stanner et al. (1997) found no difference between exposure *in utero* and exposure during infant life on various health outcomes. However, the possibility of selection bias emanating from selective mortality was not acknowledged (Rich-Edwards and Gillman 1997).

Probably the most studied historical famine is the Chinese Great Leap Forward famine of 1959-61, where scholars have attempted to address the problems surrounding scarring and selection using different sources and methods. Gørgens et al. (2012) look at growth stunting in this famine, using data from the *China Health and Nutrition Survey*. Their methodology attempts to identify scarring and selection by linking the heights of adults born before, during and after the Chinese Great Leap period in famine affected and unaffected distrincts with their own children born post-famine. The logic here is that these children only inherit the selection (genotype) and not the scarring (phenotype). They find the famine cohort was stunted by 1 to 2 cm, but that this was offset by selection of equal magnitude, leading to no overall effect on the average (Gørgens et al. 2012: Fig. 1).^4^ However, Gørgens et al. (2012, p. 103) caution that their result may be biased upwards or downwards due to the dynamics of internal migration.

Elsewhere, Kim et al. (2016) also look at the long-run health impact of the Great Leap Forward, this time using the *China Health and Retirement Longitudinal Study*. They find cohorts born during the famine years had poorer health and lower cognitive ability. They outline various selection issues (selective mortality, fertility and migration) which may influence their results, but as they are unable to address these empirically they instead suggest interpreting their findings as a lower bound of their true effects. Using the same dataset, Xu et al. (2016) attempt to see whether fetal famine exposure had a long-run health impact, measured by biomarkers and anthropometric characteristics. Their innovation is to use multiple estimation strategies which take account of selective mortality. Their primary method is a simple cohort difference, which compares the cohort born during the famine with cohorts born before and after. Using this cohort approach indicates a famine effect, but this effect disappears with their alternative estimators.^5^ They conclude this to be evidence of selective mortality; those most likely to have been affected by the famine had already died (“survival of the fittest”).

As a way to overcome sample selection biases from selective mortality in famines, scholars have attempted to test the FOH using non-famine natural experiments, such as pandemics and income shocks. Almond’s (2006) landmark study uses samples from the 1960, 1970 and 1980 US censuses to ascertain the impact of prenatal exposure to the influenza pandemic of 1918-19. He finds increased likelihood of disability, lower income and lower education of those exposed to the pandemic *in utero* (cf. Brown and Thomas 2018). Meanwhile, Banerjee et al. (2010) focus on the long-run health effects of income shocks at birth, proxied using adult male height and life expectancy. Their empirical strategy exploits the differential incidence of *phylloxera*, an insect which destroyed large segments of the French grape crop between 1863 and 1890. They use a difference-in-difference methodology to compare children born in *phylloxera*-affected areas with those born before and after *phylloxera* exposure. Every additional hectare of land used for vineyards is associated with a 3 per cent decline in height, approximately 1 cm (Banerjee et al. 2010: p. 723).

Our goal is to understand the effect of the Famine on the surviving Irish population. We therefore need to identify the potential influence that selection has on the question we pose. But we also need to think about the additional impact of selection on our underlying data source. We have identified the main sources of selection that affect our study in Table 2: selective mortality (S1), selective migration (S2), selection into crime (S3), and selection into prison (S4). S1 and S2 are different responses to famine: S1 is a biological phenomenon; S2 is socially determined.^6^ In terms of S1, the youngest and oldest are more vulnerable and comprise the majority of mortality in normal years in pre-epidemiological transition societies. Famine would exacerbate this pattern; we would expect the young and the old to be disproportionately affected by malnutrition and disease. To address the selective mortality, we control for famine intensity proxied by famine-period deaths. Meanwhile, S3 and S4 would bias results downwards as they imply only certain segments of society are represented in our data source.

**Table 2:** Competing selection effects

| Selection | Effect | Direction of bias |  |
| --- | --- | --- | --- |
|  |  | Urban sample | Rural sample |
| S1 | Selective mortality | – | + |
| S2 | Selective migration | – | + |
| S3 | Selection into crime | – | – |
| S4 | Selection into prison | – | – |

It is possible to identify the areas of the island and the demographic groups that were most affected by the Famine in terms of mortality (S1). Evidence from archaeological studies provide indications as to the selective nature of famine mortality conditions in Ireland. The main archaeological evidence comes from the excavation of the Kilkenny Workhouse burial ground, 33 km from Clonmel (Geber 2016).^7^ A high proportion of skeletal remains exhibited evidence of scurvy rather than any infectious disease, and this disproportionately affected the young.^8^ Comparisons with other burial sites show the skeletal remains in Kilkenny had a much less varied diet (Beaumont et al. 2013).^9^

However, the representativeness of the data is uncertain due to the so-called “osteological paradox”: demographic non-stationarity, selective mortality, and hidden heterogeneity in risks (Wood et al. 1992; Beaumont et al. 2015). From the perspective of our study, the most important of these is selective mortality, as osteological data can only provide information about those who died at a given age, but nothing about survivors. For example, deaths of children are selective as those most frail are more likely to die. Those who survive early life conditions are more likely to survive longer. These issues aside, the archaeological evidence suggests famine mortality was highest amongst the young, and survivors were a highly selected group.

Migration was an adaptive (and selective) response of the population to the Famine. If the poorest and weakest migrate from the affected areas, then S2 conflates with S1 and negates the impact of S1 and mortality more generally. Figure 4 plots the mortality rates against migration rates for the Famine period. While there is no obvious relationship, it does highlight the implications for our study as Dublin has net inward migration whereas Tipperary has net outward migration.

**Figure 4:**
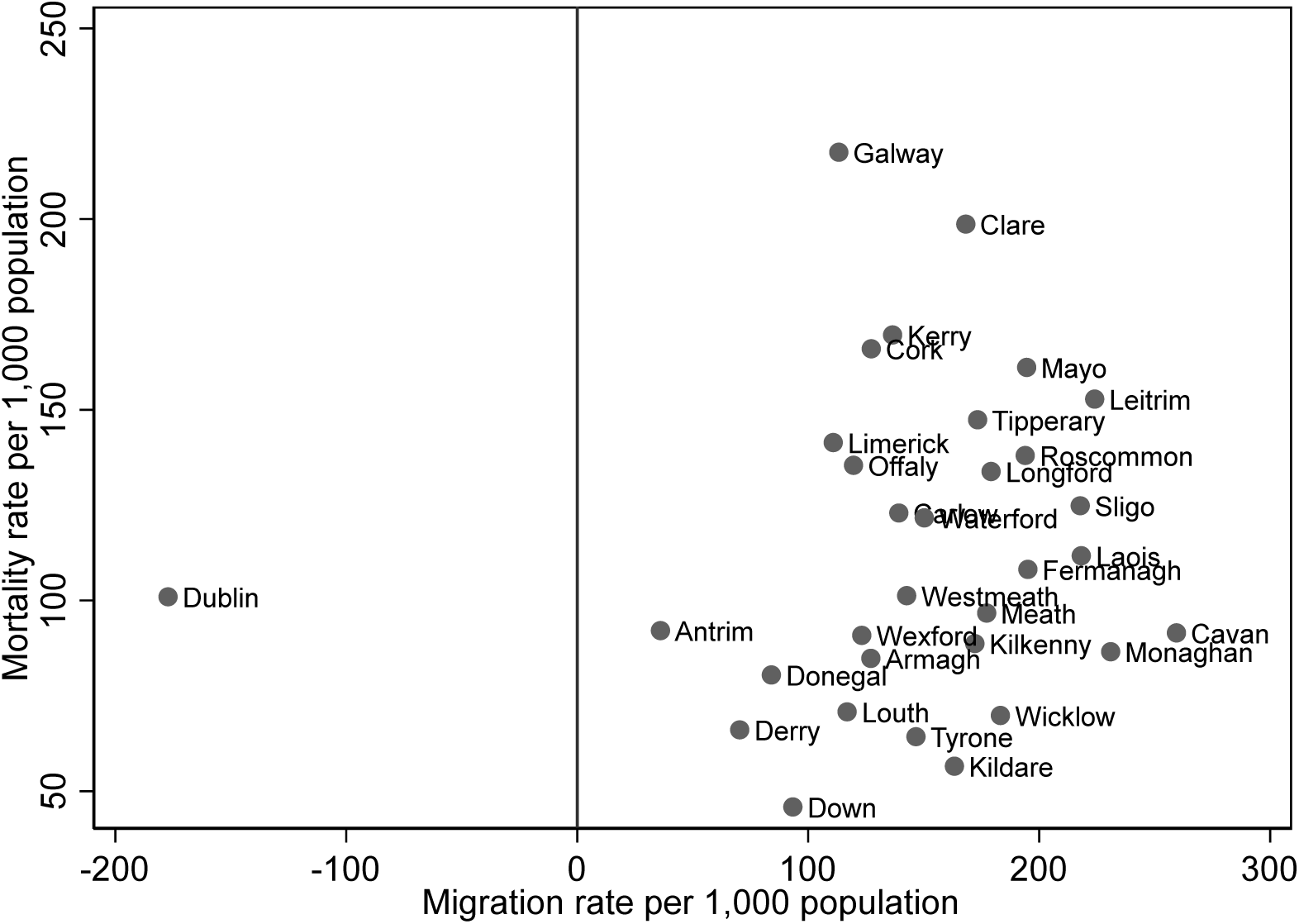
Famine mortality and famine migration *Sources:* Authors’ calculations using methodology and sources derived from Ó Gráda and O’Rourke (1997).

Understanding the composition of migrants (e.g., were they the poorest, the richest, or of median income) helps understand the direction of the selection effect. Ó Gráda and O’Rourke (1997) document that emigration was beyond the reach of the very poorest, although this poverty trap was somewhat alleviated by remittances from first-wave migrants (cf. Schrier 1958). Another outlet was internal migration. These migration flows were directed towards urban centres. In this case, migration from the heavily affected rural areas would lead to selection (under-estimating the true effect) and migration to urban areas would lead to scarring (over-estimating the true effect).

There is a wider debate ongoing in economic history on migrant selection. Recent work by Connor (2019) using US and Irish censuses finds evidence of negative selection of Irish migrations, while Collins and Zimran (2019) find intergenerational convergence of Famine-era Irish migrants to the US in terms of occupational status. More careful comparison of anthropometric datasets either side of the Atlantic is needed to tease out the full ramification of this selection – including whether international migrants hailing from Ireland’s urban and rural areas were relatively better off than their non-migrating peers. Given both the archaeological evidence and documented migration patterns, it is evident that some element of both S1 and S2 is present, but would differ depending on urban and rural compositions. Our analysis of nationally-averaged data (composing both urban and rural samples) would possibly see the effects of S1 and S2 averaged out.

As for the prison data that we use to test the effect of the Famine, there are issues of selection into crime (S3) and then selection into prison (S4). Effectively the selection issue here relates to the representativeness of prisoners. This has recently become a key concern in the anthropometrics literature (Bodenhorn et al. 2017): were prison inmates representative of the general population, a criminal underclass, or selecting into crime differently over time? McLaughlin et al. (2021) present and discuss the various selection issues surrounding the prison data used in the present study. They find any changes in selection into crime across time were more likely to have been the result of institutional (changes in criminal classification) rather than economic factors (various business cycle indicators), but do find that there is a relationship between the two during the period of the Famine.

Our strategy to address S3 is to “sample” data from the prison population incarcerated only *after* the Famine concluded, thereby excluding the selected population that can distort our isolation of a famine effect. In terms of S4, it could be possible that specific crimes and the people who commit such offences are more likely to be represented in our sample. We control for the type of crime committed in our analysis to address this concern. We find a disproportionate number of those incarcerated were not doing time for serious crimes, but were rather incarcerated for short spells for the social offence of “drunkenness”. Given the nature of this crime, which only began to be enforced in the 1870s (McLaughlin et al. 2021), we argue that drunkards are likely to be broadly representative of the wider population of Ireland.

We test the impact of selection effects S3 and S4 more rigorously by comparing the observable attributes of the prison population with the corresponding census waves and re-weight our estimates in accordance with how our sample deviates from census observables. Appendices to this present paper include econometric specifications in which we attempt to further address the issues surrounding the representativeness of prisoners. These robustness exercises do not alter our conclusions.

## 4. Data

To gain any insight into the health conditions of the Irish population requires the use of an archival source of data in which stature was consistently recorded for cohorts born before, during and after the Famine. In terms of generating new evidence, our contribution is the use of anthropometric information extracted from one such source: prison registers. These unique and rich historical documents facilitate a direct inference of the impact of famine-related nutritional and dietary change. We sample cohorts born before, during and after the Famine from the complete population of prisoners incarcerated for any period of time between 1854 (after the Famine had ended) and 1910 in two locations in Ireland: Dublin and Clonmel (locations indicated in Figure 3).^10^ These registers provide detailed information on every prisoner who entered the prison system on categories including their height, age, crime, location of birth and residence, occupation, religion and level of literacy.^11^ The data were recorded by prison administrators and are standardised in nature; new inmates were required to undergo an obligatory medical examination by a prison doctor (Breathnach 2014). Our data do not constitute only hardened criminals; “drunkenness” was criminalised and punishable with a custodial sentence.^12^

Today, stature is widely used as an anthropometric indicator of the health status of a population in various literatures (e.g., Fogel 2004; Floud et al. 2011; Deaton 2013). Heights reflect health and living standards from an outcome-oriented viewpoint. The mean stature of a population is a function of both official and unofficial income, including subsistence farming and public goods provisioning (Steckel 1995). Heights are also sensitive to income inequality (Deaton 2008). The height of a generation is determined around the time of its birth, with health standards of mothers as well as nutritional and health standards of infants playing a crucial role (Eveleth and Tanner 1976; Steckel 1995).

Deaton (2007) argues cross-country evidence on height is difficult to interpret. His case in point is Africa, where the population is the world’s poorest but also the tallest of all developing regions. He disregards genetics as an explanation in the African case, suggesting instead selection may be greater there than scarring. He also highlights the variation in heights across social groups within poor countries as evidence against a genetic explanation. Similarly, in our study we disregard genetic determinants as we are focusing on the population of one specific island which shares a common “gene pool”. An additional advantage is our sample is drawn from a single social group: the least successful in Irish society, those selecting or selected into the criminal population and subsequently caught, convicted and incarcerated as punishment for their crimes.

Over the period of our study, there were three “prison regimes” (following McLaughlin et al. 2021). The first, from 1791 to 1853, saw the removal of a segment of criminals from the UK as a punishment for what were deemed the most severe crimes: over this period, 26,500 convicts were transported to Australia (Kilcommins et al. 2004: p. 17).^13^ Transportation was replaced with penal servitude and this was administrative change occurred in 1877, when prisons throughout Ireland were centralised under the General Prisons Board. This resulted in a change in the classification of prisoners being held in Kilmainham and Clonmel: they were to be used for ‘untried and prisoners under sentence, males for sentences not exceeding 12 months and females not exceeding 6 months’ (BPP 1878-79, p. 37). From 1877 onwards, prisoners serving longer-term sentences were sent to specialist convict prisons: Lusk (Dublin), Mountjoy (Dublin), and Spike Island (Cork). McLaughlin et al. (2021) conclude there is little evidence of selection into crime across variables other than these institutional changes.

The personal characteristics used in our study include height, age, year of birth, literacy status, religion and nature of crime committed.^14^ Our key variable used to identify any possible famine effect is average adult height. For Kilmainham Gaol, mean male height, after excluding children and adolescents below the age of 17, is 168.1 cm (see Table 3 for summary statistics). Height in Clonmel is somewhat higher, at 170.0 cm. This reflects generally superior biological living standards in rural Ireland.^15^ The average year of birth is 1850, with sufficient variation to capture living standards before, during and after the Famine. At point of incarceration, males were on average aged 33.1 years in Kilmainham and 38.5 years in Clonmel. For Clonmel Gaol Protestants make up 1.7 per cent of the population, while for Kilmainham Gaol this is 9.4 per cent, reflecting the larger share of Protestants in the Greater Dublin area. The data provide for each individual their ability, separately, to read and write; we classify individuals who are able to read and write as “literate”, and “illiterate” otherwise.^16^ We find inmates of Kilmainham Gaol were generally more likely to be literate than their rural counterparts.

**Table 3:**
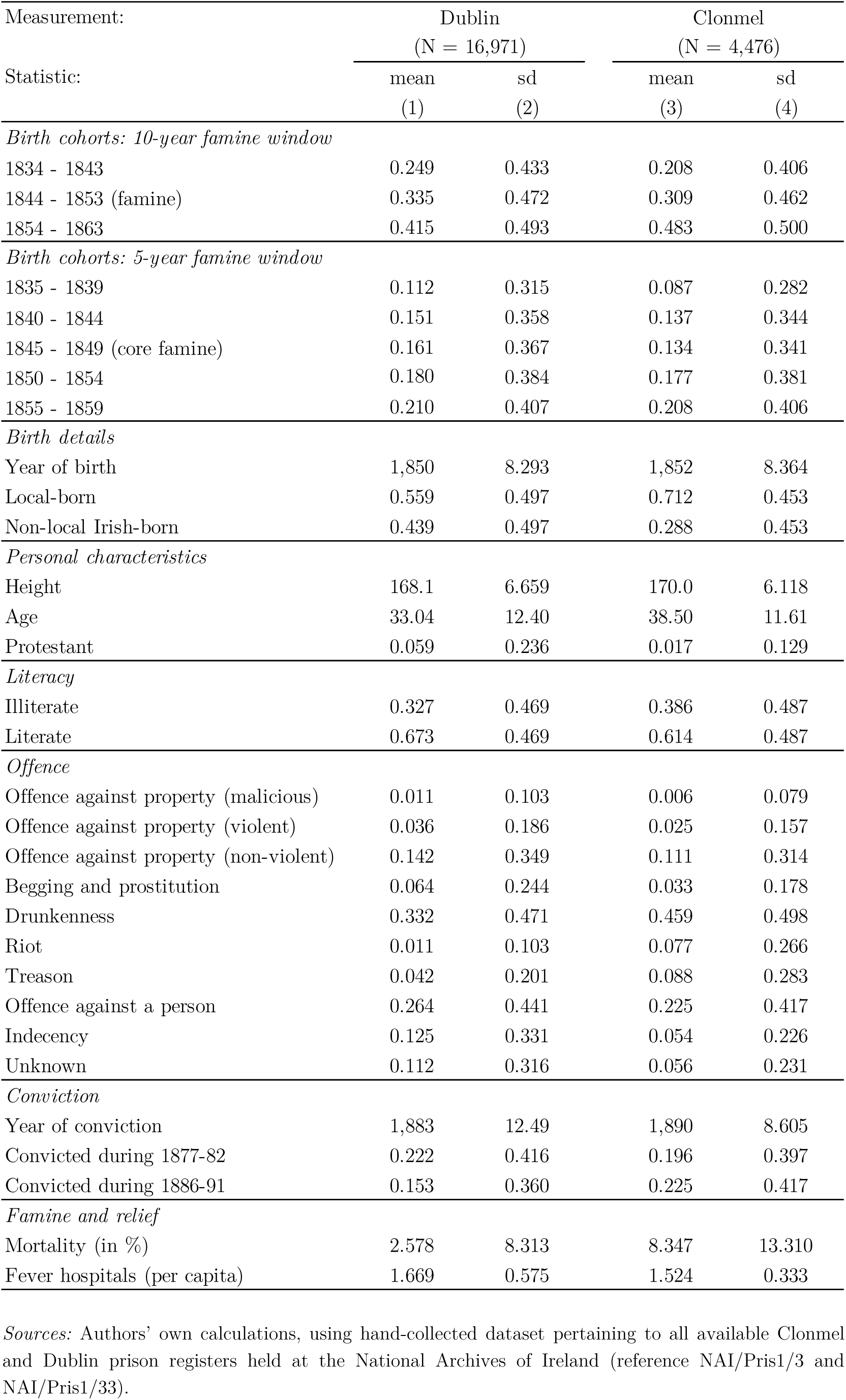
Descriptive statistics

| Measurement: | Dublin<br>(N = 16,971) |  | Clonmel<br>(N = 4,476) |  |
| --- | --- | --- | --- | --- |
| Statistic: | mean | sd | mean | sd |
|  | (1) | (2) | (3) | (4) |
| *Birth cohorts: 10-year famine window* |  |  |  |  |
| 1834 - 1843 | 0.249 | 0.433 | 0.208 | 0.406 |
| 1844 - 1853 (famine) | 0.335 | 0.472 | 0.309 | 0.462 |
| 1854 - 1863 | 0.415 | 0.493 | 0.483 | 0.500 |
| *Birth cohorts: 5-year famine window* |  |  |  |  |
| 1835 - 1839 | 0.112 | 0.315 | 0.087 | 0.282 |
| 1840 - 1844 | 0.151 | 0.358 | 0.137 | 0.344 |
| 1845 - 1849 (core famine) | 0.161 | 0.367 | 0.134 | 0.341 |
| 1850 - 1854 | 0.180 | 0.384 | 0.177 | 0.381 |
| 1855 - 1859 | 0.210 | 0.407 | 0.208 | 0.406 |
| *Birth details* |  |  |  |  |
| Year of birth | 1,850 | 8.293 | 1,852 | 8.364 |
| Local-born | 0.559 | 0.497 | 0.712 | 0.453 |
| Non-local Irish-born | 0.439 | 0.497 | 0.288 | 0.453 |
| *Personal characteristics* |  |  |  |  |
| Height | 168.1 | 6.659 | 170.0 | 6.118 |
| Age | 33.04 | 12.40 | 38.50 | 11.61 |
| Protestant | 0.059 | 0.236 | 0.017 | 0.129 |
| *Literacy* |  |  |  |  |
| Illiterate | 0.327 | 0.469 | 0.386 | 0.487 |
| Literate | 0.673 | 0.469 | 0.614 | 0.487 |
| *Offence* |  |  |  |  |
| Offence against property (malicious) | 0.011 | 0.103 | 0.006 | 0.079 |
| Offence against property (violent) | 0.036 | 0.186 | 0.025 | 0.157 |
| Offence against property (non-violent) | 0.142 | 0.349 | 0.111 | 0.314 |
| Begging and prostitution | 0.064 | 0.244 | 0.033 | 0.178 |
| Drunkenness | 0.332 | 0.471 | 0.459 | 0.498 |
| Riot | 0.011 | 0.103 | 0.077 | 0.266 |
| Treason | 0.042 | 0.201 | 0.088 | 0.283 |
| Offence against a person | 0.264 | 0.441 | 0.225 | 0.417 |
| Indecency | 0.125 | 0.331 | 0.054 | 0.226 |
| Unknown | 0.112 | 0.316 | 0.056 | 0.231 |
| *Conviction* |  |  |  |  |
| Year of conviction | 1,883 | 12.49 | 1,890 | 8.605 |
| Convicted during 1877-82 | 0.222 | 0.416 | 0.196 | 0.397 |
| Convicted during 1886-91 | 0.153 | 0.360 | 0.225 | 0.417 |
| *Famine and relief* |  |  |  |  |
| Mortality (in %) | 2.578 | 8.313 | 8.347 | 13.310 |
| Fever hospitals (per capita) | 1.669 | 0.575 | 1.524 | 0.333 |

*Sources:* Authors' own calculations, using hand-collected dataset pertaining to all available Clonmel and Dublin prison registers held at the National Archives of Ireland (reference NAI/Pris1/3 and NAI/Pris1/33).

A set of control variables is based on the type of offence for which an individual was imprisoned, following the example of Blum et al. (2017). Offences are distinguished into seven sub-categories, adapting a contemporary criminal classification: offences against the person, violent and non-violent offences against property, begging and prostitution, drunkenness, rioting, treason and a category for offences related to indecency. Duplicate observations due to multiple imprisonment of recidivists are excluded from the analysis to avoid double counting. We categorise all prisoners according to the nature of their crime to be able to control for offence-specific effects in our analysis, reported below.^17^ Finally, the main analysis is restricted to a sub-group of individuals older than 17 years-of-age. Populations exposed to low biological standards of living usually reach their final adult height in their late-teen years – in extreme cases even later (see Eveleth and Tanner 1976). We add variables to control for any such catch-up growth in our samples.^18^

## 5. Empirical Strategy

The empirical strategy used to test for a “famine effect” relies on a test of differences in final average male height between the Famine-born cohort and the cohorts born in the decades prior to and immediately after the Famine. This strategy aims to capture the effect of malnutrition and worsening disease environment experienced by the Famine-born cohort, i.e., stunted growth. Our working hypothesis is this effect should result in a height drop for individuals in locations where famine conditions were most severe, in rural Ireland (see Figure 3). Our alternative hypothesis is we find no effect, which implies there was “extreme selection”.

We use a broad definition of the Famine. While the main famine years are 1845 to 1852, we include the birth years 1844 and 1853 to account for famine exposure during infancy (1844 birth cohort) and *in utero* (1853 birth cohort). The following formula outlines the testing framework:

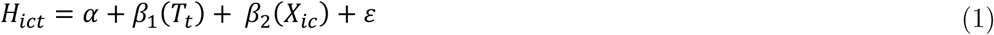

where *H* denotes height of individual *i* who was born in county *c* at time *t*. All results are conditional on a time trend which tests for differences in height between the cohorts born before, during and after the Famine, indicated by vector *T*. Other control variables, denoted by vector *X*, include literacy, religion, birth county-fixed effects, and the type of crime committed. *X* also contains controls for conviction periods when selection into the prison sample might have differed from normal times. We run our analysis separately for each prison.

We use the aforementioned framework to test for a famine effect in two different OLS settings. Our first strategy uses the rationale of a structural break to test for differences in height between a sequences of birth cohorts. This is also known as calculating the simple cohort difference. Our second strategy is based on the idea of a difference-in-difference approach, where the rate of excess mortality in individual *i*’s county during the Famine proxies famine exposure, i.e., the level of malnutrition and disease environment. In this setup, mortality rates help to distinguish high excess mortality regions of birth, such as rural Munster and Connacht, from low mortality regions, such as parts of Ulster and the Greater Dublin area.

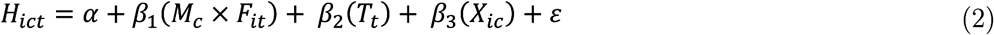

In this latter framework we keep controls for time (*T*) to capture general time-specific effects, but add an interaction term (*M*_*c*_ × *F*_*it*_) combining famine-related mortality (*M*_*c*_) and information about whether an individual was born during the Famine (*F*_*it*_). This term proxies for famine severity and is equal to the excess mortality (in per cent) in an individual’s birth county if this individual was born during the Famine, and zero otherwise. To obtain a total effect in this setup, we combine all mortality-related effects obtained in the regression analyses with the general height trend estimated using birth cohort dummies. In models which solely use birth period effects, this combined metric is equal to the difference in height between pre-Famine and Famine-born cohorts. For models based on a combined strategy of using birth period dummies together with excess mortality, we compute the aggregate effect by entering the observed excess mortality into Formula 2.

OLS may be inefficient if the errors are non-normal. Implicitly, conventional estimators test for an average famine effect which applies to all points on the height distribution equally (see Figure 5). However, it is reasonable to assume the Famine did not affect tall and short individuals equally. Indeed, elsewhere scholars have found short individuals tend to come from lower socioeconomic backgrounds, while for tall individuals the opposite is true (Steckel 1995). We address the presumption selective mortality and migration (S1 and S2 above) disproportionately affected some parts of the height distribution in two ways: by using a Tobit specification, and by conducting a quantile regression analysis.

**Figure 5:**
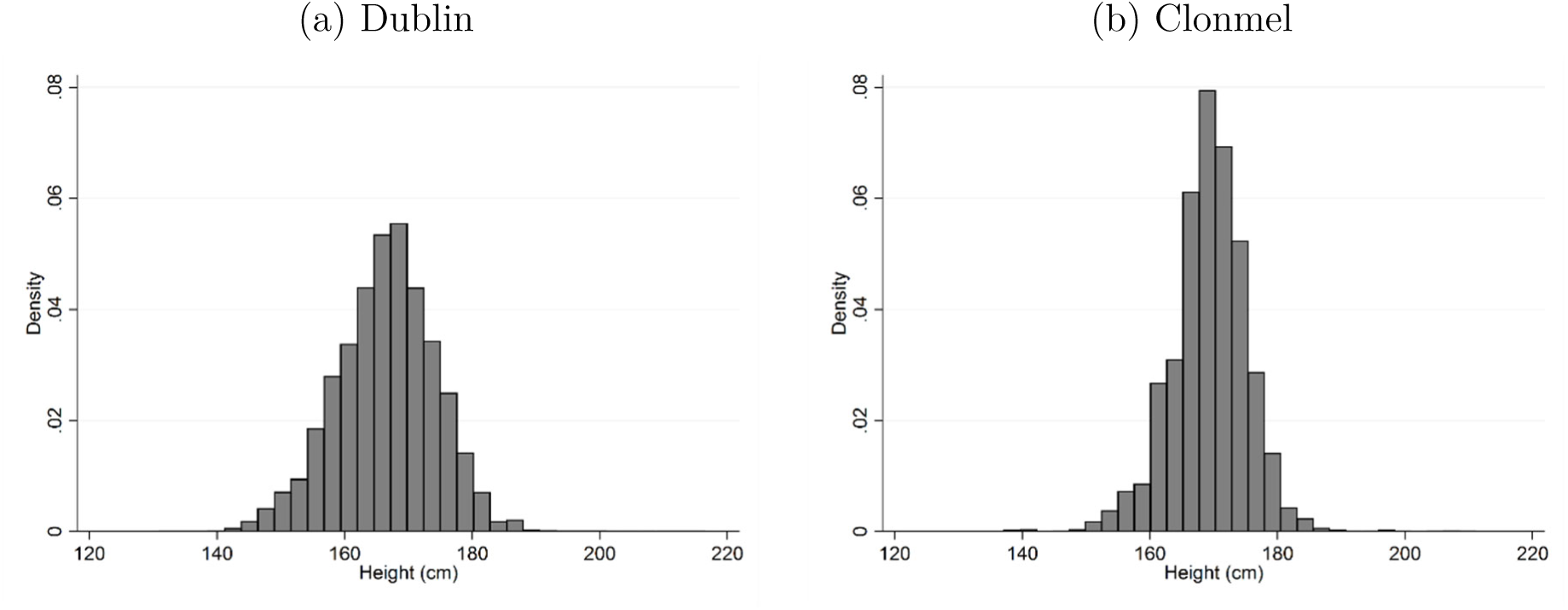
Height histograms, male prisoners *Sources:* Authors’ calculations, using Clonmel and Dublin prison registers.

Tobit regression techniques address potential selection bias in the left-hand-side of the height distribution, choosing a cut-off point which is below, but close, to the analysed samples’ median height. If the Tobit specification leads to substantially different results compared to the OLS results, we can conclude selective mortality and migration disproportionately affects the left hand-side of height distribution. We use Formula 4, assuming the specification of individual height outlined in Formula 3.

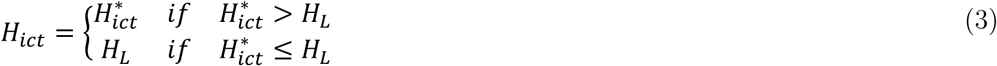

where *H*_*L*_ is censored from below, and 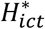is a latent variable specified as:

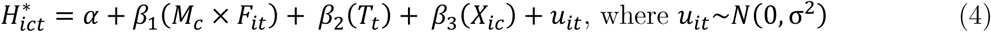

Quantile regression is more robust to non-normal errors and outliers. This method allows us to consider the impact of a set of covariates on the entire height distribution.^19^ We use a set of quantile regression models in a 10-year famine window setting to test for differences in the magnitude of the Famine within the height distributions. The conditional quantile function is defined as the *τ*-th quantile *Q*_*Y*|X_ *τ*|*x* of the conditional distribution function *F*_*Y*|X_ *y*|*x* for the dependent variable height *y* given the explanatory variable *x*, is a binary variable identifying a Famine-born individual. We report results for Dublin and County Tipperary using the following quantiles to assess differences in Famine exposure of different height percentiles: *τ* = 10; *τ* = 25 ;*τ*_3_ = 50; *τ*_4_ = 75; and *τ*_5_ = 90.

## 6. Principal Results

Table 4 reports the origin and measurement of the individuals in each sample used, and separates city-born from rural-born individuals to enable a precise analysis of the effects of famine in this historical setting.^20^ Regression coefficients indicate the magnitude of the famine effect according to Formulas 1 and 2, the number of observations, and the coefficient of determination.^21^ In addition, we report famine-related mortality in the birth counties of the individuals in each sample, and the total estimated change in height due to exposure to the Famine. These results are shown on the bottom line of the table.

**Table 4:** OLS and Tobit specifications, 10-year famine window

| Measurement: | Dublin |  |  |  | Clonmel |  |  |  |
| --- | --- | --- | --- | --- | --- | --- | --- | --- |
| Sample origin: | Dublin | Ireland (excl. Leinster) |  |  | Tipperary | Ireland (excl. Munster) |  |  |
| Estimator: | OLS | OLS | OLS | Tobit | OLS | OLS | OLS | Tobit |
|  | (1) | (2) | (3) | (4) | (5) | (6) | (7) | (8) |
| Constant | 167.18***<br>(5,011.14) | 168.22***<br>(246.80) | 168.18***<br>(255.58) | 167.46***<br>(295.36) | 169.52***<br>(440.15) | 168.28***<br>(196.34) | 168.05***<br>(187.17) | 170.18***<br>(172.68) |
| *Birth period* |  |  |  |  |  |  |  |  |
| 1834 - 1843 | *reference* | *reference* | *reference* | *reference* | *reference* | *reference* | *reference* | *reference* |
| 1844 - 1853 (famine) | -0.27***<br>(-8.39) | 0.20<br>(0.69) | -0.72<br>(-0.93) | -1.04<br>(-1.42) | 0.70**<br>(2.35) | -0.05<br>(-0.04) | 0.75<br>(0.52) | 1.03<br>(1.00) |
| 1854 - 1863 | -0.67***<br>(-16.80) | 0.86<br>(1.74) | 0.86<br>(1.71) | 0.78*<br>(1.75) | 0.58**<br>(2.11) | 0.30<br>(0.27) | 0.31<br>(0.28) | 0.22<br>(0.27) |
| *Famine effect* |  |  |  |  |  |  |  |  |
| Mortality in % |  |  | 0.03<br>(1.32) | 0.04**<br>(2.13) |  |  | -0.05<br>(-0.96) | -0.02<br>(-0.65) |
| Observations | 9,485 | 2,011 | 2,015 | 2,015 | 3,318 | 460 | 460 | 460 |
| R-squared | 0.06 | 0.09 | 0.09 |  | 0.03 | 0.10 | 0.10 |  |
| *Famine mortality in area under observation (in %)* | -0.7 | 33.6 | 33.6 | 33.6 | 29.4 | 16.9 | 16.9 | 16.9 |
| *Estimated height change due to famine (in cm)* | -0.27 | 0.20 | 0.29 | 0.30 | 0.70 | -0.05 | -0.10 | 0.69 |

Notes: The dependent variable is final adult height in cm. Birth periods are binary variables identifying whether an individual was born during one of the nominated time periods. Famine effect is proxied by excess mortality during the Famine in % of the population in the area under observation; 'famine mortality in area under observation' is a weighted average of the Famine-related mortality experienced in the home counties of individuals included in each sub-sample in order to proxy for the Famine exposure of a sub-sample. Unreported control variables constitute age at measurement, time of imprisonment (prison regime dummies), religion, literacy status, county fixed effects and the type of offence. Reference category is an adult, pre-famine-born illiterate male Catholic who committed an offence against property (non-violent) and was imprisoned before 1877. Robust t-statistics in parentheses; standard errors are corrected for clustering and autocorrelation by clustering at the county level; \*\*\* p<0.01, \*\* p<0.05, \* p<0.1.

For males born in Dublin County, our results suggest a modest height decline throughout the 30-year period under observation. Height declined by 0.3 cm during the Famine, and by an additional 0.7 cm after the Famine (Table 4, Model 1). We compare this Dublin-born, Dublin-imprisoned group with another sample which consists of within-Ireland migrants. By contrast, for Irishmen born outside Leinster we do not find such an obvious effect. In Models 2 to 4, the results suggest a total famine effect of 0.2 to 0.3 cm, but these coefficients are not statistically significant. For the period after the Famine, we find the height of those born in locations closer to Dublin tended to decrease (Model 1), while height in areas most heavily affected by the Famine stagnated, or even increased (Models 2 to 4).

We use another sample, inmates of Clonmel Gaol in County Tipperary, to gain insights into height developments in rural Ireland (Table 4, Models 5 to 8). We separately assess the heights of those hailing from Tipperary and those born in areas of Ireland outside the province of Munster. As for Tipperary’s urban-born individuals, we find an increase of 0.7 cm for the Famine-born cohort, and a further increase of 0.6 cm afterwards (Model 5). While mortality figures imply a significant famine effect, there was no corresponding drop in height; indeed, combining the mortality and height coefficients implies a stagnation or even an increase in height for the Famine-born generation. If the Famine left its mark on the Irish population, we expect to find signs of stunting in this rural sample of Munster-born prisoners. But here, results suggest heights were either unaffected (Models 6 and 9), or exhibited an increase (Model 8) in height during the Famine – although these coefficients are not statistically significant. Again, this finding is consistent with selection rather than scarring.

A set of quantile regressions, reported in Table 5, help us to further illustrate the relative roles of selection and scarring. They allow us to test whether this average effect can also be observed for each denoted percentile, or, indeed, whether the famine effect varies across the height distribution. Recall that for Dublin-born individuals, our OLS estimates suggest a minimal decrease in average height (as shown again in Table 5, Model 1). The Greater Dublin area was only modestly exposed to the Famine. Mortality was modest, but a general increase in food prices may have permanently affected the health of lower classes. For the 10th percentile, we observe an increase in height by approximately 1.0 cm, suggesting selection more than outweighed scarring. At the 25th, 50th and 75th percentiles we observe a conventional famine effect in that heights dropped by 0.5 cm, 1.1 cm and 0.4 cm. Interestingly, heights decreased more at the median than at the 25th or 75th percentile. We interpret this as evidence of the changing role of selection and scarring across the distribution. We do not observe a statistically significant change in height at the 90th percentile, suggesting the Famine had an effect which was minimal or absent among this class.

**Table 5:** OLS and quantile specifications, 10-year famine window

| Measurement: | Dublin |  |  |  |  |  | Clonmel |  |  |  |  |  |
| --- | --- | --- | --- | --- | --- | --- | --- | --- | --- | --- | --- | --- |
| Sample origin: | Dublin |  |  |  |  |  | Tipperary |  |  |  |  |  |
| Estimator / percentile: | OLS | 10 | 25 | 50 | 75 | 90 | OLS | 10 | 25 | 50 | 75 | 90 |
|  | (1) | (2) | (3) | (4) | (5) | (6) | (7) | (8) | (9) | (10) | (11) | (12) |
| Constant | 167.18***<br>(672.12) | 157.48***<br>(300.64) | 162.88***<br>(642.48) | 168.13***<br>(522.98) | 171.64***<br>(328.13) | 175.90***<br>(414.83) | 169.52***<br>(440.15) | 162.03***<br>(173.24) | 166.81***<br>(359.63) | 169.87***<br>(374.76) | 172.38***<br>(345.79) | 175.58***<br>(318.68) |
| *Birth period* |  |  |  |  |  |  |  |  |  |  |  |  |
| 1834 - 1843 | *reference* | *reference* | *reference* | *reference* | *reference* | *reference* | *reference* | *reference* | *reference* | *reference* | *reference* | *reference* |
| 1844 - 1853 (famine) | -0.27<br>(-1.40) | 0.95***<br>(2.61) | -0.48<br>(-1.62) | -1.10***<br>(-4.67) | -0.44*<br>(-1.78) | -0.32<br>(-0.82) | 0.70**<br>(2.35) | 1.27<br>(1.15) | 0.83**<br>(2.37) | 0.31<br>(0.85) | 0.96***<br>(2.85) | 0.95<br>(1.62) |
| 1854 - 1863 | -0.67***<br>(-3.35) | 0.63<br>(1.43) | -0.48**<br>(-1.99) | -1.15***<br>(-5.22) | -1.08***<br>(-5.49) | -1.11***<br>(-2.64) | 0.58**<br>(2.11) | 0.64<br>(0.57) | 0.83**<br>(2.12) | -0.00<br>(-0.00) | 0.96**<br>(2.41) | 0.95<br>(1.54) |
| Observations | 9,485 | 9,485 | 9,485 | 9,485 | 9,485 | 9,485 | 3,318 | 3,318 | 3,318 | 3,318 | 3,318 | 3,318 |
| R-squared | 0.06 |  |  |  |  |  | 0.03 |  |  |  |  |  |

*Notes:* The dependent variable is final adult height in cm. Birth periods are binary variables identifying whether an individual was born during one of the nominated time periods. Unreported control variables constitute age at measurement, time of imprisonment (prison regime dummies), religion, literacy status, county fixed effects and the type of offence. Reference category is an adult, pre-famine-born illiterate male Catholic who committed an offence against property (non-violent) and was imprisoned before 1877. Robust t-statistics in parentheses; where relevant, standard errors are corrected for clustering and autocorrelation by clustering at the birth-county level; \*\*\* p<0.01, \*\* p<0.05, \* p<0.1.

Recall that results for Clonmel suggest selection more than outweighs scarring, resulting in an increase in height of the famine cohort (as shown again in Table 5, Model 7). If the effect of selection (relative to scarring) is largest in locations which were most exposed to the Famine, then it is reasonable to expect this effect to be most pronounced among low percentiles. We find this positive effect was indeed strongest when using the 10th percentile as a benchmark, where we observe a height increase of 1.3 cm for the Famine-born cohort in County Tipperary (Table 5, Model 8). Indeed, quantile regression results point towards a famine effect similar to that observed in Dublin among the lowest classes. This effect is somewhat weaker for the 25th and 75th percentiles, ranging between approximately 0.8 and 1.0 cm. We find a modest positive effect for the median and 90th percentile, but these coefficients were not statistically significant.

## 7. Additional Results

We run a series of additional analyses in order to ascertain the robustness of our findings. These are reported in the Appendices. In the first exercise, reported in Appendix A, we adopt the same identification strategy as in previous tables, but: (1) we take a variety of different sub-samples from our dataset with different characteristics; and (2) we redefine our core famine period as running from 1845 to 1849 and adopt a shorter, 5-year, observation window for our difference-in-difference analysis. Our results are consistent with our main findings.

The second exercise, reported in Appendix B, uses an instrumental variable approach to address potential omitted variable bias resulting from unobserved individual characteristics. We use the number of so-called “fever hospitals” per 10,000 families in an individual’s county of origin in the era *before* the Famine to proxy a county’s capacity to address famine fever *during* the Famine. Fever hospitals were exogenous to famine exposure and correlate with our mortality variable in the first stage of a 2SLS setup. For Dublin, IV results generally confirm earlier findings. Individuals born during the Famine in Leinster were approximately 1.0 cm shorter; results for Irish immigrants born outside the region of Leinster suggest height stagnation. While we cannot confirm height increases for the Munster-born sub-sample, IV results are in line with Tobit regressions and indicate height stagnation. In general, however, results are consistent with our earlier findings for Clonmel Gaol: the absence of scarring, suggesting extreme selection.

The third exercise, reported in Appendix C, exploits the religious affiliation and literacy status of prison inmates to disentangle whether scarring and selection may be cancelling one another out. Contrary to economic intuition, we find Catholics and illiterates in the surviving population were *less* affected by the Famine, not more, despite their somewhat lower socioeconomic status.

In the fourth exercise, described in Appendix D, we re-weight our prison sample to better reflect the attributes of the general population, taken from census summaries compiled for the British Parliament. The new results are consistent with our baseline results. The evidence presented so far not only suggests the famine effect differs by location, but that this effect varies according to socioeconomic status within each location.

The fifth robustness exercise, in Appendix E, further redefines our birth cohort window length to track year-on-year height differences. However, to do so we must remove those individuals born in round years due to a phenomenon known as “age heaping”. Again, our baseline results are confirmed by this analysis in that we find no famine effect for rural Ireland, but rather a drop in height among the urban Famine-born cohorts.

A sixth exercise, Appendix F, attempts to account for possible famine stunting experienced during adolescence. We re-run our analysis with new samples of individuals who were 3-7, 8-12 and 13-17 at the end of the Famine. Much like for infants, we find a famine adolescence effect for Dublin-born individuals, but no such effect for those born elsewhere.

Finally, the seventh exercise, Appendix G, looks for evidence of scarring amongst Irish emigrants. We use a sample extracted from Fogel et al. (1990) containing the 2,977 Union Army soldiers above the age of 19 who were born in Ireland. We then adopt the same methodology as we describe in Section 6: we run an OLS regression of height (in cm) using five-year birth cohorts. We find the same height penaly pattern among Irish-born Americans fighting in the Union Army as Dublin prisoners.

## 8. Discussion and Conclusion

We adopted a research design which allows us to isolate a large sample of individuals born during the period of Ireland’s reduced food availability, and then use anthropometric indicators as a proxy for their biological standard of living. Our results are somewhat surprising. We find a positive correlation between famine severity and the health of the populations most exposed to famine-induced hunger and disease during infancy; low famine-related mortality is associated with the most severe drops in average adult height. How can an incidence of famine have no long-run health consequences for some survivors? Our research, in which we use unique and extensive hand-collected data on populations in different geographic locations across one famine episode, suggests the answer lies in the severity of the famine under study. We argue those individuals most affected by Ireland’s nineteenth-century famine underwent an “extreme selection”; those most exposed did not themselves survive in Ireland long enough to have their measurements taken; those who died, or migrated, do not, by definition, enter our Irish based samples.

The Great Irish Famine has left an indelible mark on the Irish psyche. It remains the key watershed in Irish economic and social history. It is still invoked in political discourse. And it has undoubtedly had a lasting effect on the island’s culture. But what apparently is far less obvious is its impact on the health and wellbeing of Ireland’s surviving population, those who did not themselves succumb to starvation and disease, and who stayed on the island rather than moved abroad. Yes, the Famine permanently changed the diet of the entire island. But we find only famine survivors hailing from those parts of the country which were *least* affected by the immediate effects of famine experienced any detectable permanent health consequences. From this we surmise that the scarring effect outweighs the selection effect only in Ireland’s urban centres; mortality did not select the population as much in these areas as most people survived into adulthood, including society’s least healthy.

We attribute the mechanism underpinning the selection and scarring outcomes to the demographic features of the Famine: the geographic regions and segments of society undergoing extreme selection had higher excess mortality, greater decreases in birth rates and higher cohort depletion rates. Using insights from our quantile regressions, the Famine likely disproportionately affected the poorest elements of society, those at the low end of the height distribution, while selection was minimal among Ireland’s better-off. Selective mortality (which we refer to as S1 in Section 3) was likely compounded by selective migration (S2); archaeological evidence of Famine-burials in Ireland were disproportionately young providing evidence for S1, while Famine-born Irish buried in London, for example, suggests Ireland exported its bad health (S2). Indeed, the view that Ireland exported its bad health is supported by our analysis of Union Army records from the US Civil War, where we find that soldiers born in Ireland during the Famine experienced a 2.34 cm height penalty compared to their older compatriots (see Appendix G).

The Famine resulted in, or at the very least accelerated the process of, economic change across Ireland. Post-Famine Ireland was in many respects a very different place than Ireland before the potato was blighted. The elimination through death or migration of vast swathes of society, and the collective realisation that the reliance on a single crop can have devastating consequences, permanently changed the structure of Irish agriculture, not to mention the nation’s eating habits. Land reforms precipitated by, and even instigated as a direct result of, the Famine accelerated the process of urbanisation and helped to provide industrialising cities with cheap labour inputs. But for those who grew up in the worst-affected areas and lived long enough to tell the tale of the Great Irish Famine to the next generation, its effect on their own personal physical health was apparently relatively benign.

We recommend famine severity and its associated selective mortality effect be included more explicitly in any future studies of catastrophic risks and health. Less severe famines, like other *in utero* and early childhood health shocks, can have long-lasting scarring effects on survivors. But more severe famines, like the Great Irish Famine experienced in rural areas, can act as Malthusian catastrophes in that they eliminate society’s most vulnerable and leave behind only its fittest. Without accounting for severity, anthropometric studies of the wellbeing of the survivors of natural or man-made disasters cannot draw meaningful inferences about their long-run impact on society.

## Data Availability

Data available from the authors upon request.

## Appendices

### Appendix A: Sample Origin and Famine Widow Length

An excerpt our results are presented in the main body of this paper (Table 4). Results presented in Table A1 constitute the full set of baseline results. In addition, we run separate tests for Leinster-born individuals and others born in more distant locations imprisoned in Dublin, to further differentiate between living conditions in urban and rural locations. For these groups, we do not find any change in height between the pre-Famine and Famine-born cohorts; regression coefficients suggest hardly any difference from zero, and standard errors suggest these coefficients are not statistically significant (Table A1, Models 2 to 7). Dublin and Leinster experienced a larger height decline than more heavily-affected rural locations, consistent with a scarring explanation. Meanwhile, our results for Clonmel and Tipperary are consistent with an extreme selection effect, particularly due to the selective mortality of the very young.

In fact, height in Leinster tends to stagnate, judging from the small coefficients which are not statistically significant (Models 4 and 5). When we use mortality in an individual’s county of birth to proxy for famine severity, our results are generally confirmed (Models 3, 5 and 7). The coefficients of the mortality variable are inconsistent in magnitude and marginally insignificant; mortality has a negative effect on height in Leinster and the Greater Dublin area, but a positive effect in rural Ireland (compare Model 5 with 3 and 7).

Similarly, we do not find a decrease in heights for individuals imprisoned in Clonmel Gaol (Models 8 to 14). Using excess mortality (Models 10, 12 and 14) does not suggest famine conditions brought about a height drop since the combined effect adds to +0.53 to +0.72 cm; the combined mortality and cohort effects rather indicate height increases.

Definitions of the precise length of the Great Irish Famine vary from source to source, and not each year of the Famine left an equally devastating mark on the population. We ran a simple robustness test which uses a “core Famine” window (1845-1849) versus immediately preceding and succeeding birth cohorts. If the Famine was most intense during this core period, we might detect a famine effect in this more intense period of starvation. Results presented in Table A2, however, are consistent with our earlier baseline results presented in Table A1. While we find a height drop among Dubliners (Table A2, Models 1, 4 and 5), we cannot detect such an effect for the rural-born population (Models 2, 3, 6 and 7). Similarly, the surviving population in rural Tipperary does not show any sign of early-childhood malnutrition.

### Appendix B: Famine Mortality and Fever Hospitals

Another way to test the robustness of the relationship between excess mortality caused by famine conditions and the final average height of the Famine-born cohort is to use an instrumental variable approach. An instrument helps to address potential omitted variable bias resulting from unobserved individual characteristics. We use the number of so-called “fever hospitals” per 10,000 families (henceforth hospitals per capita) in an individual’s county of origin in the era *before* the Famine to proxy a county’s capacity to address famine fever *during* the Famine. Fever hospitals were part of the medical infrastructure of pre-Famine Ireland, established from the 1830s to prevent the increase of infectious fevers (Geary 2012).

In the first stage (Formula B1) of this 2SLS procedure, we use the per capita availability of hospitals (*K*) which each individual enjoyed in any given county of birth as an instrument to explain excess mortality, resulting in (*M*_*c*_ × *F*_*it*_)′. In the second stage, we re-estimate the relationship between excess mortality and average height (Formula B2).

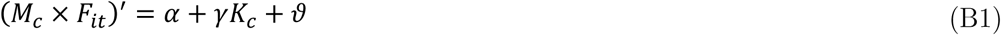

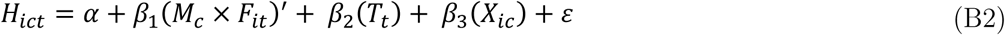

A central requirement of the exclusion restriction is the instrument must affect the left-hand-side variable only through its effect on the potentially-endogenous right-hand-side variable, i.e., excess mortality. Indeed, the history literature does not suggest a direct link between hospitals and the height of the Famine-born cohort, except through their beneficial effects on mortality and living standards during the Famine years (Geary 2012: p. 202). The presence of a hospital was not a sign of a county’s wealth; rather it was a consequence of a locality’s strong social hierarchy and the patronage of landlords, who were responsible for their funding. Fever alone accounted for almost a third of the excess mortality during the Famine (Mokyr and Ó Gráda 2002: p. 352), and so this medical infrastructure suddenly found itself at the centre of *Table A1:* OLS and difference-in-differences specifications, 10-year famine window famine relief efforts. While hospitals existed before, during and after the Famine, their role in treating fever and limiting fever-related mortality was most pronounced during the Famine.

**Table A1:**
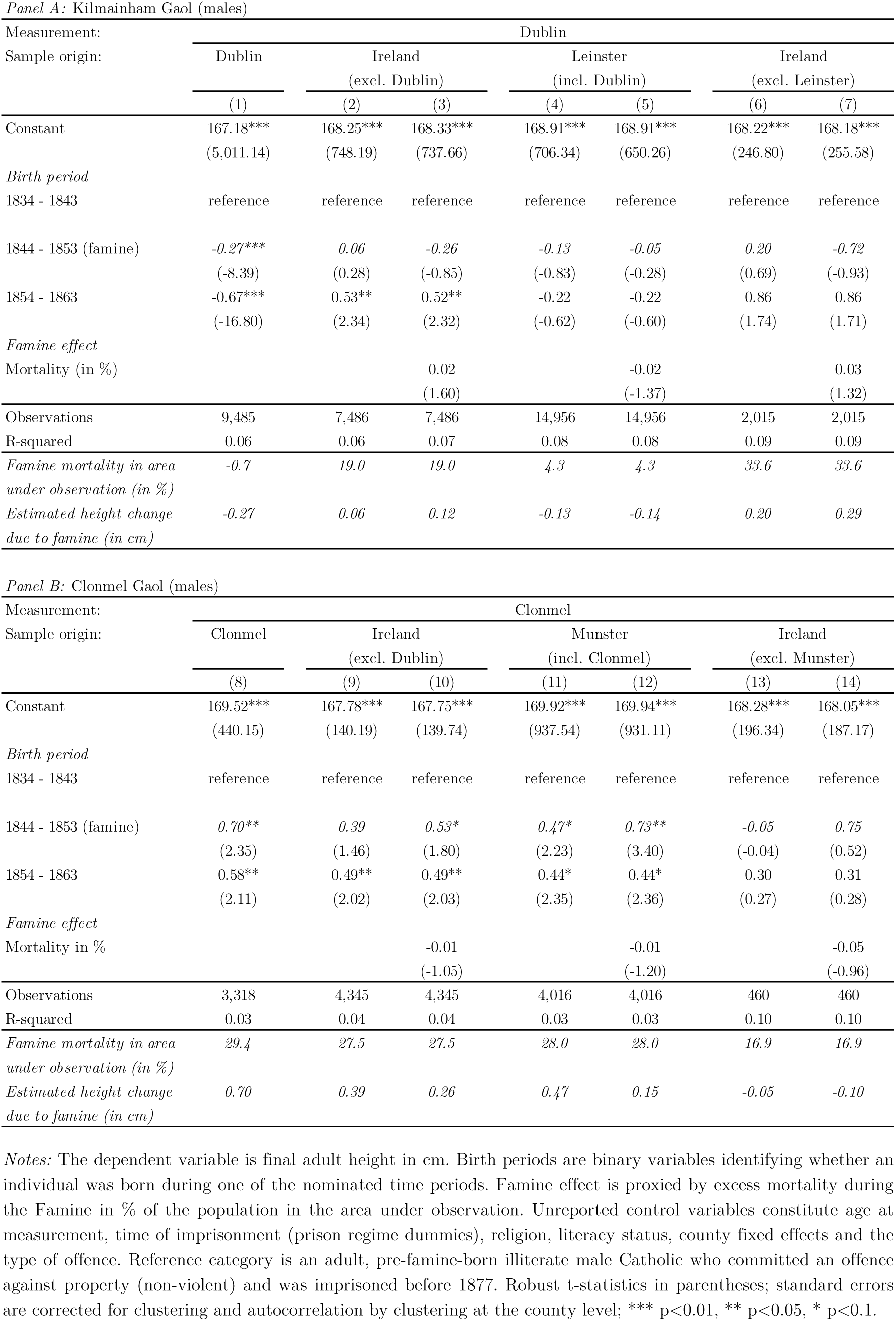
OLS and difference-in-differences specifications, 10-year famine window

**Table A2:** OLS and difference-in-differences specifications, 5-year famine window

Panel A: Kilmainham Gaol (males)
| Measurement: | Dublin |  |  |  |  |  |  |
| --- | --- | --- | --- | --- | --- | --- | --- |
| Sample origin: | Dublin | Ireland<br>(excl. Dublin) |  | Leinster<br>(incl. Dublin) |  | Ireland<br>(excl. Leinster) |  |
|  | (1) | (2) | (3) | (4) | (5) | (6) | (7) |
| Constant | 167.47***<br>(5,195.65) | 168.35***<br>(776.87) | 168.41***<br>(763.50) | 169.12***<br>(532.50) | 169.12***<br>(528.34) | 168.50***<br>(269.90) | 168.49***<br>(270.40) |
| *Birth period* |  |  |  |  |  |  |  |
| 1840 - 1844 | reference | reference | reference | reference | reference | reference | reference |
| 1845 - 1849 (core famine) | -0.69***<br>(-21.76) | -0.03<br>(-0.16) | -0.47<br>(-1.41) | -0.42**<br>(-2.15) | -0.41**<br>(-2.16) | 0.07<br>(0.19) | -0.90<br>(-1.13) |
| 1850 - 1854 | -0.46***<br>(-11.66) | -0.06<br>(-0.24) | -0.06<br>(-0.26) | -0.27<br>(-1.59) | -0.27<br>(-1.58) | 0.23<br>(0.31) | 0.23<br>(0.31) |
| 1855 - 1859 | -1.06***<br>(-29.74) | 0.61**<br>(2.51) | 0.60**<br>(2.47) | -0.46<br>(-1.04) | -0.45<br>(-1.04) | 0.98<br>(1.37) | 0.97<br>(1.35) |
| *Famine effect* |  |  |  |  |  |  |  |
| Mortality (in %) |  |  | 0.02*<br>(1.78) |  | -0.00<br>(-0.23) |  | 0.03<br>(1.68) |
| Observations | 9,467 | 7,481 | 7,481 | 14,937 | 14,937 | 2,011 | 2,011 |
| R-squared | 0.06 | 0.07 | 0.07 | 0.08 | 0.08 | 0.10 | 0.10 |
| *Famine mortality in area*<br>*under observation (in %)* | -0.7 | 19.0 | 19.0 | 4.6 | 4.6 | 33.3 | 33.3 |
| *Estimated height change*<br>*due to famine (in cm)* | -0.69 | -0.03 | -0.09 | -0.42 | -0.41 | 0.07 | 0.10 |

Panel B: Clonmel Gaol (males)
| Measurement: | Clonmel |  |  |  |  |  |  |
| --- | --- | --- | --- | --- | --- | --- | --- |
| Sample origin: | Clonmel | Ireland<br>(excl. Dublin) |  | Munster<br>(incl. Clonmel) |  | Ireland<br>(excl. Munster) |  |
|  | (8) | (9) | (10) | (11) | (12) | (13) | (14) |
| Constant | 169.69***<br>(421.66) | 170.00***<br>(864.56) | 169.99***<br>(813.30) | 167.75***<br>(542.65) | 167.69***<br>(535.84) | 167.59***<br>(135.75) | 167.53***<br>(137.24) |
| *Birth period* |  |  |  |  |  |  |  |
| 1840 - 1844 | reference | reference | reference | reference | reference | reference | reference |
| 1845 - 1849 (core famine) | 0.35<br>(0.89) | 0.16<br>(0.64) | 0.06<br>(0.10) | 0.17<br>(0.69) | 0.70<br>(0.83) | 0.56<br>(0.37) | 1.60<br>(1.32) |
| 1850 - 1854 | 0.85**<br>(2.36) | 0.85***<br>(6.86) | 0.85***<br>(6.86) | 0.76***<br>(4.20) | 0.76***<br>(4.20) | 0.39<br>(0.29) | 0.37<br>(0.28) |
| 1855 - 1859 | 0.67**<br>(2.00) | 0.62***<br>(7.21) | 0.62***<br>(7.20) | 0.77***<br>(5.30) | 0.77***<br>(5.31) | 1.13<br>(1.16) | 1.08<br>(1.11) |
| *Famine effect* |  |  |  |  |  |  |  |
| Mortality in % |  |  | 0.00<br>(0.16) |  | -0.02<br>(-0.65) |  | -0.06<br>(-1.02) |
| Observations | 3,318 | 4,016 | 4,016 | 4,345 | 4,345 | 460 | 460 |
| R-squared | 0.03 | 0.04 | 0.04 | 0.04 | 0.04 | 0.10 | 0.10 |
| *Famine mortality in areas*<br>*under observation (in %)* | 29.4 | 27.4 | 27.4 | 27.7 | 27.7 | 17.2 | 17.2 |
| *Estimated height change*<br>*due to famine (in cm)* | 0.35 | 0.16 | 0.06 | 0.17 | 0.15 | 0.56 | 0.57 |

Notes: The dependent variable is final adult height in cm. Birth periods are binary variables identifying whether an individual was born during one of the nominated time periods. Famine effect is proxied by excess mortality during the Famine in % of the population in the area under observation. Unreported control variables constitute age at measurement, time of imprisonment (prison regime dummies), religion, literacy status, county fixed effects and the type of offence. Reference category is an adult, pre-famine-born illiterate male Catholic who committed an offence against property (non-violent) and was imprisoned before 1877. Robust t-statistics in parentheses; standard errors are corrected for clustering and autocorrelation by clustering at the county level; \*\*\* p<0.01, \*\* p<0.05, \* p<0.1.

Fever hospitals were distributed highly unevenly, with the south of the island having a larger number. We consider this another piece of evidence these hospitals were not a sign of economic development or living standards in general; if hospitals had been related to income or wealth, we would expect the typical east-west division in terms of economic development observable in historical Ireland, not the north-south division visible for hospitals (Ó Gráda 1994).

We run these tests separately for males from Leinster (the province in which Dublin is located) and rural Ireland. Geary (2012) reports “famine fever” affected all parts of society, regardless of income and wealth. However, malnutrition was most severe in rural Ireland and among the poor; society’s most physically weak were more vulnerable during the Famine and were more likely to die, potentially leading to positively-skewed height distributions.^22^

The first stage results in the IV specification confirm a relationship between fever hospitals and mortality (Table B1.A). Mortality is systematically lower where hospitals were more abundant. Apart from Model 12, where there is little variation in famine-related mortality, F-statistics generally suggest this instrument is strong enough to be used in this setting. For Kilmainham Gaol, IV results suggest males born in Leinster during the Famine were only marginally shorter than their pre-famine born peers (Table B1.A, Model 3). Results for those born outside Leinster are not statistically significant, and the combined effect (regardless of standard errors) suggest height stagnation during the Famine (Models 6 and 9).

We applied the same methodology to the Clonmel sample; we report baseline OLS results alongside IV results for three different sub-sets of the data (Table B1.B). For our Munster-born and Ireland-born sub-samples, the baseline OLS results suggest an increase in height during the Famine, IV results indicate stagnation (Models 12, 15 and 18).

**Table B1:** Instrumental variable specifications, 10-year famine window

| *Panel A: Kilmainham Gaol (males)* |  |  |  |  |  |  |  |  |  |
| --- | --- | --- | --- | --- | --- | --- | --- | --- | --- |
| Measurement: | Dublin |  |  |  |  |  |  |  |  |
| Sample origin: | Leinster (incl. Dublin) |  |  | Ireland (excl. Leinster) |  |  | Ireland |  |  |
| Estimator: | OLS | IV (1st st.) | IV (2nd st.) | OLS | IV (1st st.) | IV (2nd st.) | OLS | IV (1st st.) | IV (2nd st.) |
| Dependent variable: | Height | Mortality | Height | Height | Mortality | Height | Height | Mortality | Height |
|  | (1) | (2) | (3) | (4) | (5) | (6) | (7) | (8) | (9) |
| Constant | 168.91***<br>(650.26) | -0.37<br>(-0.38) | 166.85***<br>(535.48) | 168.18***<br>(255.58) | 4.11**<br>(1.93) | 166.82***<br>(324.04) | 168.78***<br>(605.15) | 2.52<br>(2.20) | 166.74***<br>(1,069.55) |
| *Birth period* |  |  |  |  |  |  |  |  |  |
| 1834 - 1843 | reference |  | reference | reference |  | reference | reference |  | reference |
| 1844 - 1853 (famine) | -0.05<br>(-0.28) |  | -0.95<br>(-1.47) | -0.72<br>(-0.93) |  | 0.51<br>(0.52) | -0.04<br>(-0.24) |  | -0.85**<br>(-2.39) |
| 1854 - 1863 | -0.22<br>(-0.60) |  | -0.39<br>(-1.24) | 0.86<br>(1.71) |  | 0.92*<br>(1.86) | -0.09<br>(-0.23) |  | -0.26<br>(-0.79) |
| *Famine effect* |  |  |  |  |  |  |  |  |  |
| Mortality (in %) | -0.02<br>(-1.37) |  | 0.16<br>(1.48) | 0.03<br>(1.36) |  | -0.01<br>(-0.19) | 0.05***<br>(3.78) |  | 0.09*<br>(5.82) |
| Fever hospitals (per capita) |  | -1.44***<br>(-2.74) |  |  | -7.71***<br>(-3.36) |  |  | -3.80***<br>(-3.83) |  |
| Observations | 14,956 | 14,956 | 14,956 | 2,015 | 2,015 | 2,015 | 16,971 | 16,971 | 16,971 |
| R-squared | 0.08 |  | 0.07 | 0.09 |  | 0.09 | 0.09 |  | 0.07 |
| Kleibergen-Paap Wald rk |  |  | 6.8 |  |  | 12.0 |  |  | 14.8 |
| F-statistic |  |  |  |  |  |  |  |  |  |
| *Famine mortality in area under observation (in %)* | 4.6 | 4.6 | 4.6 | 33.3 | 33.3 | 33.3 | 8.2 | 8.2 | 8.2 |
| *Estimated height change due to famine (in cm)* | -0.05 |  | -0.14 | -0.72 |  | 0.39 | -0.04 |  | 0.33 |

| *Panel B: Clonmel Gaol (males)* |  |  |  |  |  |  |  |  |  |
| --- | --- | --- | --- | --- | --- | --- | --- | --- | --- |
| Measurement: | Clonmel |  |  |  |  |  |  |  |  |
| Sample origin: | Munster (incl. Clonmel) |  |  | Ireland (excl. Munster) |  |  | Ireland |  |  |
| Estimator: | OLS | IV (1st st.) | IV (2nd st.) | OLS | IV (1st st.) | IV (2nd st.) | OLS | IV (1st st.) | IV (2nd st.) |
| Dependent variable: | Height | Mortality | Height | Height | Mortality | Height | Height | Mortality | Height |
|  | (10) | (11) | (12) | (13) | (14) | (15) | (16) | (17) | (18) |
| Constant | 168.94***<br>(386.98) | -3.21<br>(5.11) | 169.23***<br>(144.15) | 169.26***<br>(174.23) | 4.55**<br>(2.31) | 169.04***<br>(183.74) | 168.70***<br>(355.79) | 0.95<br>(3.19) | 168.30***<br>(166.93) |
| *Birth period* |  |  |  |  |  |  |  |  |  |
| 1834 - 1843 | reference |  | reference | reference |  | reference | reference |  | reference |
| 1844 - 1853 (famine) | -0.47<br>(-1.38) |  | -10.8<br>(-0.79) | 0.56<br>(-0.43) |  | 2.07<br>(-0.98) | 0.19<br>(0.30) |  | 4.77<br>(-0.84) |
| 1854 - 1863 | 0.44*<br>(-2.32) |  | 0.45**<br>(-2.42) | 0.16<br>(-0.15) |  | 0.15<br>(-0.14) | 0.42**<br>(2.33) |  | 0.43**<br>(-2.38) |
| *Famine effect* |  |  |  |  |  |  |  |  |  |
| Mortality (in %) | 0.03**<br>(2.47) |  | 0.4<br>(-0.82) | -0.04<br>(-0.84) |  | -0.12<br>(-0.84) | 0.01<br>(0.37) |  | -0.16<br>(-0.77) |
| Fever hospitals (per capita) |  | 2.73<br>(0.66) |  |  | -3.74<br>(-2.94) |  |  | -2.84<br>(1.09) |  |
| Observations | 4,016 | 4,016 | 4,016 | 460 | 460 | 460 | 4,476 | 4,476 | 4,476 |
| R-squared | 0.03 |  |  | 0.02 |  |  | 0.02 |  |  |
| Kleibergen-Paap Wald rk |  |  | 0.4 |  |  | 8.7 |  |  | 6.8 |
| F-statistic |  |  |  |  |  |  |  |  |  |
| *Famine mortality in area under observation (in %)* | 28.1 | 28.1 | 28.1 | 16.0 | 16.0 | 16.0 | 26.8 | 26.8 | 26.8 |
| *Estimated height change due to famine (in cm)* | 0.37 |  | 0.60 | -0.08 |  | 0.15 | 0.46 |  | 0.48 |

*Notes:* The dependent variable is final adult height in cm. Birth periods are binary variables identifying whether an individual was born during one of the nominated time periods. Famine effect is proxied by excess mortality during the Famine in % of the population in the area under observation, and instrumented using the per capita availability of fever hospitals prior to the Famine. Unreported control variables constitute age at measurement, time of imprisonment (prison regime dummies), religion, literacy status, county fixed effects and the type of offence. Reference category is an adult, pre-famine-born illiterate male Catholic who committed an offence against property (non-violent) and was imprisoned before 1877. This IV strategy works well for samples that provide sufficient variation in the exogenous and potentially endogenous variable, such as in the case of the Kilmainham sample; the Clonmel sample provides less geographical variation and accordingly F-statistics remain somewhat below the threshold of 10. Robust t-statistics in parentheses; where relevant, standard errors are corrected for clustering and autocorrelation by clustering at the birth-county level; \*\*\* p<0.01, \*\* p<0.05, \* p<0.1.

### Appendix C: Religion, Literacy and Famine Vulnerability

We perform a robustness check by testing for a famine effect among the most vulnerable elements of Irish society: Catholics and illiterates. In the pre-Famine period, most of the Irish population was Roman Catholic and in turn Catholics made up the bulk of poorest in society, those most likely to have been exposed to famine conditions. Historically, Catholics were discriminated following the Reformation, and this only intensified after the Glorious Revolution, from 1688.^23^ This discrimination was felt throughout all levels of Catholic society.^24^

With regards to illiteracy, there is good empirical evidence literacy and stature are correlated. Short individuals and individuals with inferior human capital are disadvantaged in official labour markets, while for taller individuals the opposite is true.^25^ If illiteracy and human height have a negatively association, and famine exposure and human height are also negatively related, then illiteracy and famine exposure may similarly be negatively related.

The evidence we review in this study suggests scarring and selection may have opposite effects in severe famines and so may even cancel one another out. This may prevent us from detecting the true magnitude of the *in utero* and early-childhood impact of the Great Irish Famine. By using interaction variables which combine information on time of birth as well as religion and illiteracy, we test for a difference in famine exposure between Protestants and Catholics, and between literates and illiterates. If scarring and selection are of a similar magnitude, but in opposite directions, then the height of Protestants and illiterates may reveal a stronger famine effect if their superior social status allowed these strata to attenuate the effects of famine-induced malnutrition and disease. In other words, if reduced famine exposure brought about limited scarring compared to Catholics and illiterates, we may infer the observed height of Protestants and literates reflects a stronger famine effect.

We use a testing framework which is similar to the one in our main analysis (see Formulas 1 and 2). Formula C1 illustrates our approach:

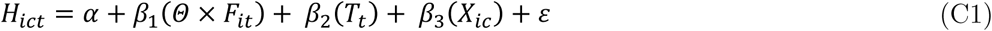

where *H*_*ict*_ is a function of a vector *T*, which tests for differences in height between the cohorts born before, during and after the Famine. Again, vector *X* includes controls for literacy, religion, county-fixed effects, and the type of crime committed. *X* also contains controls for conviction periods when selection into the prison sample might have differed from normal times. Most importantly, *Θ* is a parameter which can be used to interact famine exposure (*F*) with either Catholicism or illiteracy.

Results of this exercise are reported in Table C1. When we test for differences by religion, the effects presented in Models 1 to 4 refer to Famine-born Catholic Dubliners. Our hypothesis suggests more selection (relative to scarring) of this societal group should lead the interaction variable identifying Famine-born Catholics to take on a positive coefficient. Indeed, the corresponding effects shown in Model 1 suggest there was a general negative famine effect for Dublin-born individuals, leading to a height drop of approximately 2.5 cm. Catholics, however, are characterised by less scarring, and more selection, leading to a somewhat less pronounced height drop of 1.9 cm. Catholics in general are shorter than their Protestant peers – consistent with the literature. And so despite their inferior status in the Irish society, Catholics’ height was *less* affected by the Famine compared to Protestants’ heights.

Recall that in the general regression setup we did not distinguish between religious groups with the interaction term. Our new results in Table C1 suggest a weaker famine effect for the Catholic majority and the stronger famine effect for the Protestant community to some extent cancelled one another out, therefore explaining why we did not originally detect a famine effect. This explanation only holds for Dublin- and Leinster-born individuals, however: we ran the same test on rural-Irish born individuals, but failed to detect such an effect; the baseline variable proxying a general famine effect, nor the interaction terms, reflect a difference in famine effect for Catholics (see Models 3 and 4, as well as Models 9 to 12).

Similarly, we use literacy as the distinctive feature to test for differences in famine effect between literates and illiterates. Again, our hypothesis suggests illiterate members of society should have experienced more selection, relative to scarring, leading to a somewhat weaker observed famine effect than for literate individuals. The effects for Dublin- and Leinster-born individuals suggest a general negative effect for the Famine-born cohort (Models 5 and 6). While we expected illiterates to be more vulnerable than literates, the interaction effect for Dublin-born individuals suggests Famine-born illiterates’ famine effect is positive, and reflects somewhat *less* exposure to famine conditions compared with literates. Again, we are of the opinion these height differences among Dublin-born individuals reflect differences in selection, where illiterates experienced more selection than scarring relative to literate individuals. We find ambiguous effects for the rural all-Ireland samples (Models 7 and 8); here we find neither a general famine effect, nor a consistent and economically or statistically significant effect for illiterates.

**Table C1:** Difference-in-differences specifications with interaction effects, 5-year famine window

|  |  |  |  |  |  |  |  |  |
| --- | --- | --- | --- | --- | --- | --- | --- | --- |
| *Panel A: Kilmainham Gaol (males)* |  |  |  |  |  |  |  |  |
| Measurement: | Dublin |  |  |  | Dublin |  |  |  |
| Sample origin: | Dublin | Leinster | Ireland<br>(w/o Dublin) | Ireland<br>(w/o Leinster) | Dublin | Leinster | Ireland-born<br>(w/o Dublin) | Ireland<br>(w/o Leinster) |
|  | (1) | (2) | (3) | (4) | (5) | (6) | (7) | (8) |
| Constant | 168.61***<br>(1,837.43) | 169.92***<br>(324.09) | 168.78***<br>(415.98) | 169.28***<br>(222.52) | 168.29***<br>(2,342.55) | 169.71***<br>(362.36) | 168.68***<br>(432.59) | 169.27***<br>(231.26) |
| *Birth period* |  |  |  |  |  |  |  |  |
| 1834 - 1843 | reference | reference | reference | reference | reference | reference | reference | reference |
| 1845 - 1849 (core famine) | -2.49***<br>(-23.28) | -1.81***<br>(-4.14) | -0.61<br>(-1.37) | -0.10<br>(-0.08) | -0.79***<br>(-19.66) | -0.60***<br>(-3.65) | -0.04<br>(-0.11) | 0.79<br>(1.76) |
| 1850 - 1855 | -0.49***<br>(-12.02) | -0.28<br>(-1.60) | -0.07<br>(-0.31) | 0.18<br>(0.26) | -0.50***<br>(-12.39) | -0.29<br>(-1.63) | -0.07<br>(-0.31) | 0.17<br>(0.24) |
| *Religion and literacy* |  |  |  |  |  |  |  |  |
| Catholic | -0.65***<br>(-6.97) | -0.16<br>(-0.46) | 0.63<br>(1.69) | 0.72<br>(1.06) | -0.30***<br>(-4.14) | 0.11<br>(0.36) | 0.73**<br>(2.07) | 0.66<br>(1.17) |
| Illiterate | -0.49***<br>(-32.95) | -0.65***<br>(-4.81) | -1.05***<br>(-4.51) | -1.47*<br>(-1.80) | -0.51***<br>(-43.68) | -0.72***<br>(-4.53) | -1.05***<br>(-4.55) | -1.03<br>(-1.21) |
| *Interaction effects* |  |  |  |  |  |  |  |  |
| Famine-born * Catholic | 1.88***<br>(16.73) | 1.45***<br>(5.02) | 0.61<br>(1.31) | 0.21<br>(0.15) |  |  |  |  |
| Famine-born * Illiterate |  |  |  |  | 0.16***<br>(2.96) | 0.42<br>(1.63) | -0.03<br>(-0.04) | -3.15***<br>(-3.32) |
| Observations | 9,522 | 15,017 | 7,518 | 2,023 | 9,522 | 15,017 | 7,518 | 2,023 |
| R-squared | 0.06 | 0.08 | 0.06 | 0.10 | 0.06 | 0.08 | 0.06 | 0.10 |
| *Panel B: Clonmel Gaol (males)* |  |  |  |  |  |  |  |  |
| Measurement: | Males (Clonmel) |  |  |  | Males (Clonmel) |  |  |  |
| Sample origin: | Tipperary | Munster | Ireland<br>(w/o Dublin) | Ireland<br>(w/o Munster) | Tipperary | Munster | Ireland<br>(w/o Dublin) | Ireland<br>(w/o Munster) |
|  | (9) | (10) | (11) | (12) | (13) | (14) | (15) | (16) |
| Constant | 169.14***<br>(170.71) | 169.97***<br>(277.42) | 168.37***<br>(218.86) | 168.17***<br>(126.23) | 169.36***<br>(169.91) | 170.13***<br>(323.93) | 168.50***<br>(240.85) | 167.94***<br>(113.01) |
| *Birth period* |  |  |  |  |  |  |  |  |
| 1834 - 1843 | reference | reference | reference | reference | reference | reference | reference | reference |
| 1845 - 1849 (core famine) | 2.03<br>(0.51) | 1.22<br>(1.36) | 0.57<br>(0.39) | -0.38<br>(-0.16) | -0.09<br>(-0.18) | -0.36<br>(-1.13) | -0.34<br>(-1.05) | 0.76<br>(0.40) |
| 1850 - 1855 | 0.85**<br>(2.36) | 0.85***<br>(6.87) | 0.76***<br>(4.20) | 0.40<br>(0.30) | 0.82**<br>(2.29) | 0.82***<br>(6.47) | 0.73***<br>(4.05) | 0.38<br>(0.29) |
| *Religion and literacy* |  |  |  |  |  |  |  |  |
| Catholic | 1.54*<br>(1.71) | 0.86<br>(1.23) | 0.14<br>(0.15) | -1.05<br>(-0.93) | 1.39<br>(1.54) | 0.78<br>(1.32) | 0.11<br>(0.12) | -0.82<br>(-0.73) |
| Illiterate | -0.99***<br>(-4.50) | -0.83***<br>(-5.34) | -0.76***<br>(-4.09) | 0.43<br>(0.56) | -1.12***<br>(-4.77) | -0.99***<br>(-8.55) | -0.92***<br>(-6.08) | 0.50<br>(0.72) |
| *Interaction effects* |  |  |  |  |  |  |  |  |
| Famine-born * Catholic | -1.69<br>(-0.42) | -1.08<br>(-1.58) | -0.41<br>(-0.30) | 1.05<br>(0.35) |  |  |  |  |
| Famine-born * Illiterate |  |  |  |  | 0.95<br>(1.46) | 1.13***<br>(4.03) | 1.11***<br>(3.50) | -0.49<br>(-0.22) |
| Observations | 3,318 | 4,016 | 4,345 | 460 | 3,318 | 4,016 | 4,345 | 460 |
| R-squared | 0.03 | 0.04 | 0.04 | 0.10 | 0.03 | 0.04 | 0.04 | 0.10 |

*Notes:* The dependent variable is final adult height in cm. Birth periods are binary variables identifying whether an individual was born during one of the nominated time periods. Unreported control variables include age at measurement, time of imprisonment (prison regime dummies), county fixed effects and the type of offence. Reference category is an adult, pre-famine-born illiterate male Catholic who committed an offence against property (non-violent) and was imprisoned before 1877. Interaction effects between Famine-born and Catholic tests the difference in famine exposure between Protestants and Catholics. Interaction effects between Famine-born and illiteracy tests the difference in famine exposure between literate and illiterate individuals. Standard errors are corrected for clustering and autocorrelation by clustering at the county level; \*\*\* p<0.01, \*\* p<0.05, \* p<0.1.

### Appendix D: Overrepresentation and Weighted Regressions

In our main findings, we use a “targeted” sample in that prisoners constitute the exact individuals which we would expect to have been most affected by famine: the poor and the working classes.^26^ However, this targeting may bias our results: perhaps society’s more successful individuals, who survived the Famine but are underrepresented in our prison samples, were most affected by famine conditions in terms of scarring. We attempt to discern whether the overrepresentation of the poor and working classes in the prison population affects our results, and whether a more representative imprisonment of Irishmen would potentially have led to different conclusions.

To achieve this, we compare data from prison populations with data from four census waves – 1871, 1881, 1891 and 1901 – to assess the degree of selection in the prison population across two dimensions: religion and literacy. We find Catholics and illiterate individuals are overrepresented in our prison population. In Appendix C we found Catholics and illiterates were less likely to show a famine effect due to their increased vulnerability and their increased exposure to selection relative to scarring. If we use weights to mimic the religious and educational composition of the whole of Irish society, we may also adjust the relative influence of scarring and selection effects, potentially leading to a different study outcome.

In practical terms, then, we use the shares of Catholics and illiterates reported in the census record for the counties of Dublin and Tipperary, and compare them with population shares in corresponding prison population. Our methodology follows the rationale of survey research, where usually response rates differ between various population groups and must be reweighted before inferences can be drawn. Table D1 shows the share of illiterate inhabitants of Dublin according to the census data for 1871, 1881, 1891 and 1901. In 1871, for example, 25 per cent of Catholics in Dublin reported to be illiterate, whereas 33 per cent of the corresponding Catholic prison population was illiterate. We find this pattern across all four census records for Catholics, but not for Protestants. Protestants are generally more literate than Catholics, and illiteracy rates in Protestant census and prison populations do not differ substantially.

We use the shares presented in Table D1 to create adjustment weights, which we use in a regression framework. Formula D1 illustrates how we compute a representation rate, *θ*, which is the illiteracy rate in the prison population (*n*) and the illiteracy rate in the general population according to the census (*N*). Adjustment weights are obtained by computing the inverse of *θ* (formula D2). Accordingly, adjustment weights to correct for illiteracy are near the value of one for Protestants, reflecting a minimal misrepresentation in the prison population. For Catholics, adjustment weights reflect a stronger misrepresentation compared to the general population.

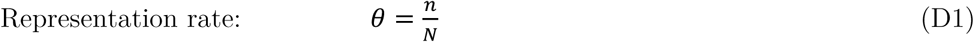

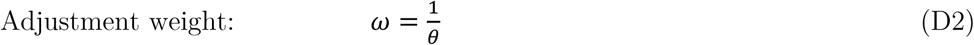

We also found the relative share of Protestants and Catholics differs between census population and prison population. In 1871, for example, 71 per cent of the population in Dublin was reported to be Catholic, whereas 90 per cent of the prison population reported to be Catholic, suggesting overrepresentation of Catholics. We compute adjustment weights using the methodology outlined above. Adjustment weights for Catholics are below one to adjust for overrepresentation in the prison population, whereas weights for Protestants aim at increasing their relative weight.

By contrast, the composition of the Clonmel Gaol population is more representative than that of Kilmainham Gaol (not shown here). Weights for the period 1871 to 1901 aiming to adjust the relative shares of literate and illiterate inmates also suggest the most overrepresented population group is illiterate Catholics, resulting in a weight below the value of one. Illiterate Protestants and literate population groups in general are fairly close to a weight of one, suggesting over and underrepresentation is only modest. Similarly, we do not find a strong overrepresentation of Catholics in Clonmel Gaol during this period, resulting in adjustment values close to the value of one. The final adjustment weights, combining both literacy and religion, which are computed using aforementioned methodology, are reported in Table D2.

We re-run our basic regressions setup where we test for a change in height between the pre-Famine cohort (born 1840-44) and the Famine-born cohort. We present results for unweighted regression, i.e., where no adjustment weights are used, alongside the corresponding regression model which uses weights, adjusting for over and underrepresentation specified in Table D3. By and large, results of both sets of regressions, for Dublin and Clonmel, are robust to the inclusion of our weights. As for inmates of Kilmainham Gaol, no noteworthy changes are detected for Dublin- or rural-born inmates. For the sub-sample of Leinster-born individuals, which includes also the Greater Dublin area, weighted regressions result in lower standard errors and a slight increase in effect size, suggesting a modest famine effect of similar magnitude as in the urban-only sub-sample. Weighted regressions for Clonmel result in somewhat lower coefficients and higher standard errors. Nevertheless, we still do not find evidence of a famine effect for Tipperary-born and rural-born Irish (excl. Munster).

**Table D1:** Illiteracy and religious composition of census and prison populations, Dublin

| Year | Illiteracy |  |  |  | Religious composition |  |  |  |
| --- | --- | --- | --- | --- | --- | --- | --- | --- |
|  | Catholics |  | Protestants |  | Catholics |  | Protestants |  |
|  | Census | Prison | Census | Prison | Census | Prison | Census | Prison |
| 1901 | 0.11 | 0.27 | 0.11 | 0.11 | 0.71 | 0.93 | 0.29 | 0.07 |
| 1891 | 0.24 | 0.34 | 0.12 | 0.10 | 0.70 | 0.94 | 0.30 | 0.06 |
| 1881 | 0.27 | 0.35 | 0.08 | 0.12 | 0.72 | 0.91 | 0.28 | 0.09 |
| 1871 | 0.25 | 0.33 | 0.16 | 0.17 | 0.71 | 0.90 | 0.29 | 0.10 |

**Table D2:** Final adjustment weights, Dublin and Tipperary

| Year | Dublin |  | Tipperary |  |
| --- | --- | --- | --- | --- |
|  | Catholics | Protestants | Catholics | Protestants |
| 1901 | 0.76 | 4.14 | 0.99 | 1.17 |
| 1891 | 0.74 | 5.00 | 0.98 | 1.50 |
| 1881 | 0.79 | 3.11 | 0.97 | 2.00 |
| 1871 | 0.79 | 2.90 | 0.98 | 1.40 |

### Appendix E: Year-on-Year Analysis and Age Heaping

The principal empirical strategy we use in our analysis is to compare the average height of birth cohorts before, during and after the Famine, which we define in either 10-year or 5-year windows. We choose these multi-year windows partly because we cannot be entirely confident about the accuracy of the birth year which newly-incarcerated prisoners provide to prison officials on entry to the prison system. This is due to “age heaping”, a well-documented phenomenon where individuals who are uncertain of their true age round their age statements to multiples of five or ten. In this appendix, we employ an alternative sampling strategy which allows us to be more precise about the year of birth, and so enables us to determine whether our principal results are driven by the choice of cohort window. In short, we remove all individuals from our sample which report a round age, enabling us to report year-on-year height differences. Unfortunately, this alternative strategy raises additional issues of representativeness, which we discuss below.

Age data often display excess frequencies at round or attractive ages, such as even numbers and multiples of five, leading to heaped distributions. A society’s propensity to age heap has been linked directly to its overall level of human capital (Mokyr and Ó Gráda 1982); trends in age heaping reveal trends in human capital accumulation (A’Hearn et al. 2009). Blum et al. (2017) find significant amount of age heaping among Ireland’s prison population, but find this disappears by the end of the nineteenth century. Figure E1 demonstrates there is also heaping in the sample of prisoners which we take for our analysis.

**Table D3:** OLS and weighted specifications, 5-year famine window

| *Panel A: Kilmainham Gaol (males)* |  |  |  |  |  |  |  |  |
| --- | --- | --- | --- | --- | --- | --- | --- | --- |
| Measurement: | Dublin |  |  |  |  |  |  |  |
| Sample origin: | Dublin |  | Leinster |  | Ireland (excl. Dublin) |  | Ireland (excl. Leinster) |  |
| Estimator: | OLS | Weighted | OLS | Weighted | OLS | Weighted | OLS | Weighted |
|  | (1) | (2) | (3) | (4) | (5) | (6) | (7) | (8) |
| Constant | 167.56***<br>(6,533.35) | 167.40***<br>(5,302.48) | 169.18***<br>(484.82) | 169.04***<br>(1,007.80) | 168.32***<br>(535.74) | 168.30***<br>(384.24) | 168.27***<br>(221.75) | 167.64***<br>(186.41) |
| *Birth period* |  |  |  |  |  |  |  |  |
| 1840 - 1844 | *reference* | *reference* | *reference* | *reference* | *reference* | *reference* | *reference* | *reference* |
| 1845 - 1849 (core famine) | -0.83***<br>(-28.74) | -0.88***<br>(-23.91) | -0.45<br>(-1.66) | -0.68***<br>(-4.18) | 0.18<br>(0.71) | -0.11<br>(-0.37) | 0.39<br>(0.89) | 0.54<br>(0.99) |
| 1850 - 1854 | -0.60***<br>(-18.65) | -0.20***<br>(-3.05) | -0.30<br>(-1.28) | -0.18<br>(-1.58) | 0.03<br>(0.12) | -0.06<br>(-0.21) | 0.25<br>(0.39) | 0.63<br>(1.13) |
| Observations | 7,882 | 7,882 | 12,238 | 12,238 | 6,006 | 6,006 | 1,650 | 1,650 |
| R-squared | 0.07 | 0.06 | 0.09 | 0.09 | 0.08 | 0.08 | 0.11 | 0.11 |
| *Panel B: Clonmel Gaol (males)* |  |  |  |  |  |  |  |  |
| Measurement: | Clonmel |  |  |  |  |  |  |  |
| Sample origin: | Tipperary |  | Munster |  | Ireland (excl. Dublin) |  | Ireland (excl. Munster) |  |
| Weights: | OLS | Weighted | OLS | Weighted | OLS | Weighted | OLS | Weighted |
|  | (9) | (10) | (11) | (12) | (13) | (14) | (15) | (16) |
| Constant | 169.87***<br>(386.46) | 169.84***<br>(373.48) | 169.88***<br>(1,642.06) | 169.56***<br>(1,475.60) | 166.37***<br>(584.85) | 165.95***<br>(709.40) | 165.74***<br>(93.70) | 165.85***<br>(105.99) |
| *Birth period* |  |  |  |  |  |  |  |  |
| 1840 - 1844 | *reference* | *reference* | *reference* | *reference* | *reference* | *reference* | *reference* | *reference* |
| 1845 - 1849 (core famine) | 0.61<br>(1.40) | 0.29<br>(0.62) | 0.36<br>(1.27) | 0.01<br>(0.02) | 0.35<br>(1.15) | -0.06<br>(-0.18) | 0.51<br>(0.27) | 0.05<br>(0.02) |
| 1850 - 1854 | 0.84**<br>(2.18) | 0.78**<br>(1.97) | 0.87***<br>(11.66) | 0.79***<br>(9.21) | 0.79***<br>(4.63) | 0.62***<br>(2.92) | 0.48<br>(0.30) | -0.10<br>(-0.06) |
| Observations | 2,739 | 2,739 | 3,308 | 3,308 | 3,565 | 3,565 | 370 | 370 |
| R-squared | 0.04 | 0.04 | 0.05 | 0.05 | 0.05 | 0.06 | 0.11 | 0.13 |

*Notes:* The dependent variable is final adult height in cm. Birth periods are binary variables identifying whether an individual was born during one of the nominated time periods. Unreported control variables constitute age at measurement, time of imprisonment (prison regime dummies), religion, literacy status, county fixed effects and the type of offence. Reference category is an adult, pre-famine-born illiterate male Catholic who committed an offence against property (non-violent) and was imprisoned before 1877. OLS regressions are identical to those reported in Table 4. Weighted regressions use weights taken from Table D2. Robust t-statistics in parentheses; standard errors are corrected for clustering and autocorrelation by clustering at the county level; \*\*\* p<0.01, \*\* p<0.05, \* p<0.1.

Removing all those prisoners who give an age ending either with a five or ten means that we can be more confident about capturing only those individuals who knew precisely in which year they were born. This enables us to compare individuals born in years of the Famine which were severely affected in terms of excess mortality, such as 1847, with other years which were affected to a much lesser extent, such as 1845.^27^ However, removing these individuals from our sample creates two further selection issues: (1) some removed individuals were likely reporting their age accurately, and just happened to have a rounded age at the time of their incarceration; and (2) by removing those prisoners reporting a rounded age, we may be failing to capture those individuals who display low levels of human capital. Both selection issues are likely to work in opposite directions, and it is difficult to determine what the overall selection effect is.

**Figure E1:**
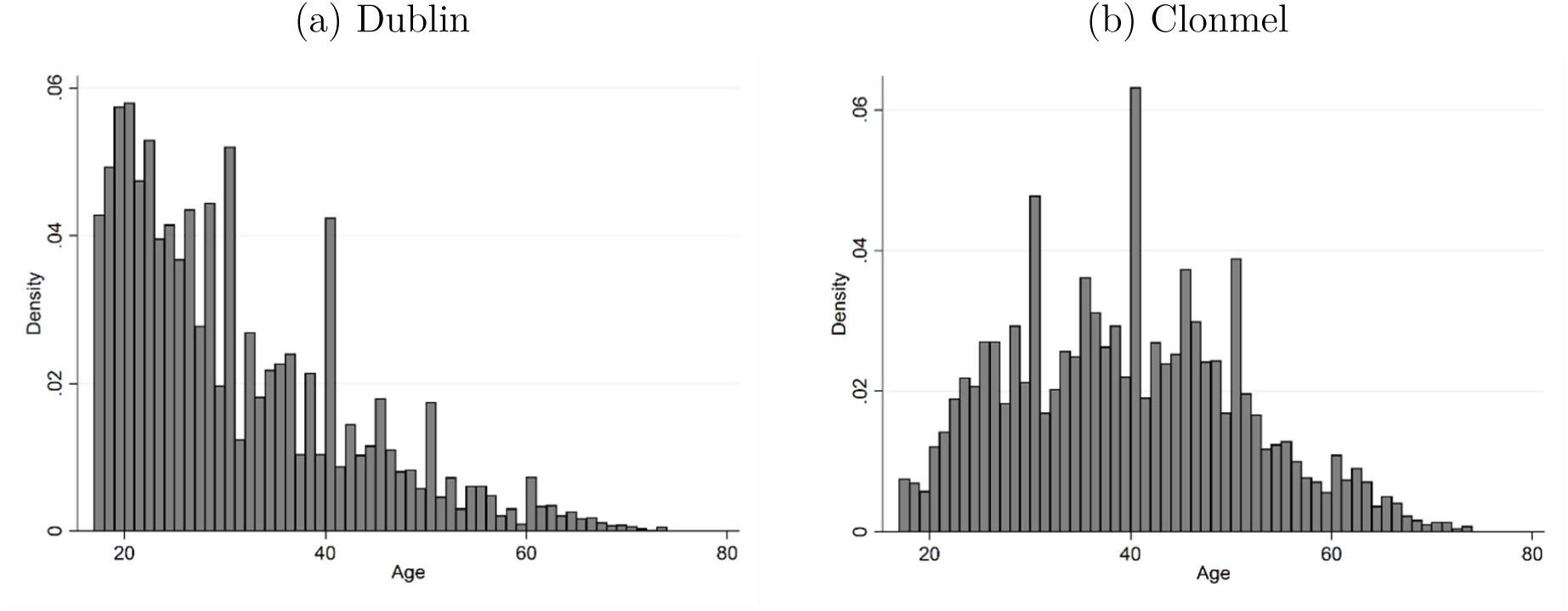
Age histograms, male prisoners *Sources:* Authors’ calculations, using Clonmel and Dublin prison registers.

**Figure E2:**
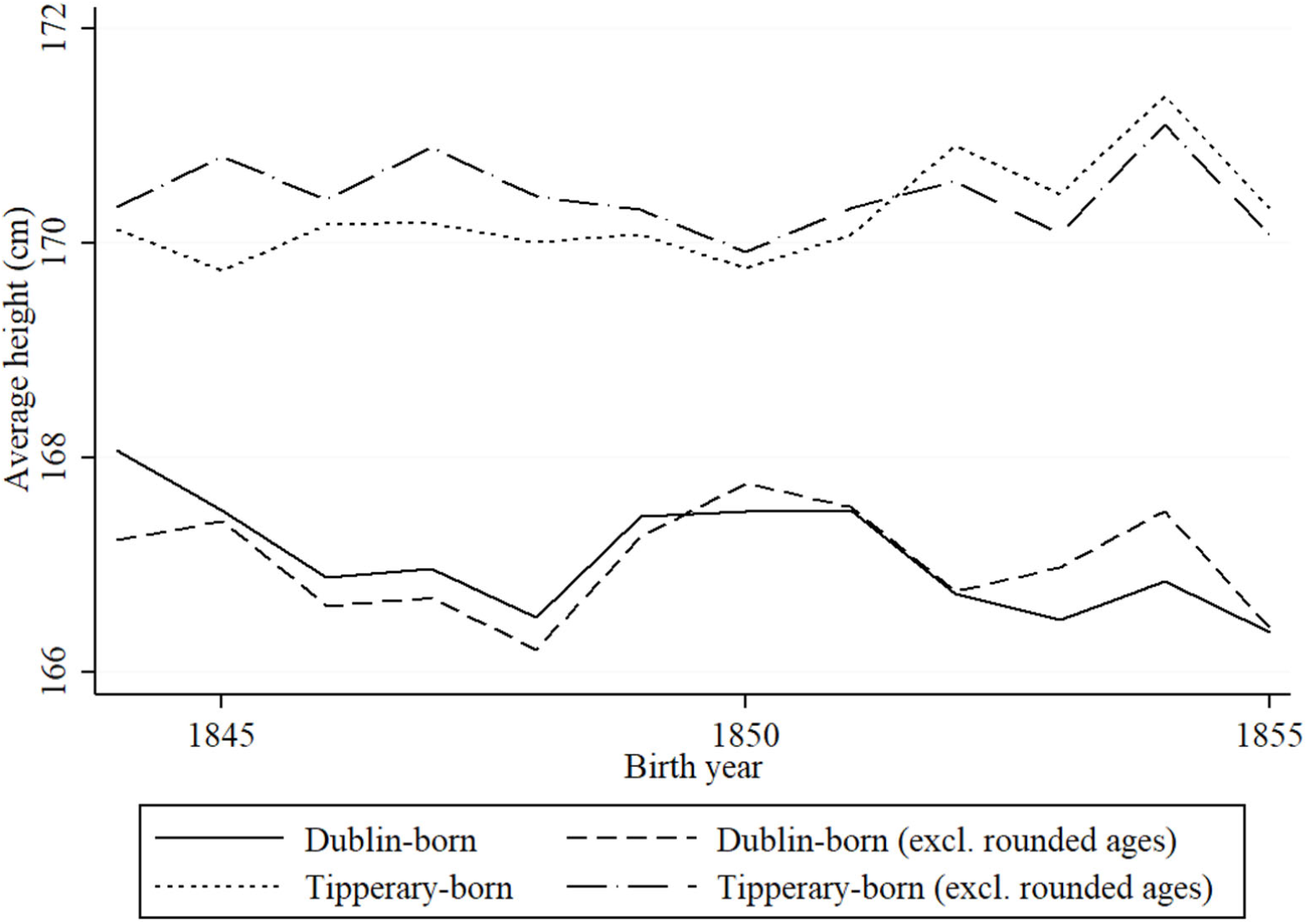
Height trends in rural and urban Ireland, 1844-55 *Sources:* Authors’ calculations, using Clonmel and Dublin prison registers.

Figure E2 displays the results of our year-on-year analysis graphically.^28^ Each height trend was computed from the coefficients of the year of birth and the regression model’s intercept. We plot height trends of all individuals in our samples alongside the subset of individuals who reported non-rounded ages. One of the most striking feature of the figure is the so-called “urban height penalty”, the consequence of poorer diet and sanitation in the city of Dublin. In terms of the aims of our study, and in common with our earlier analysis, we find no significant impact of famine on the heights of those born during the Famine period among Tipperary-born prisoners, but find a difference of approximately 1.0 cm for those born in Dublin. Removing those individuals born in rounded years does not change the height trend or the magnitude of these results.

### Appendix F: Famine Exposure and Adolescent Growth

The principal goal of our analysis has been to measure whether there was an *in utero* impact of the Great Irish Famine on terminal adult height. The motivation for this goal has been the FOH scholarship, which establishes the health and disease environment of a child’s mother from its inception to about age two are the most important determinants of adult stature.

But early infancy is not the only period in which human beings experience rapid growth in stature; teenagers also experience rapid growth. Indeed, as illustrated by the growth velocity curve in Figure F1, the second-most important period of growth is adolescence, historically between 12 and 16 for well-nourished boys (Komlos et al. 2009). Typically, teenage years are presented in the historical anthropometrics literature as a period in which those undernourished in infancy experience catch-up growth (Steckel 1986; Schneider 2017). However, if nutritional and disease environments are impaired during this period, this catch-up may never take place. Moradi (2010) shows heights of adolescents in sub-Saharan Africa stagnated at the same time as countries experienced negative shocks GDP between the 1950s and 1980s. More pertinent still to our own study, Depauw and Oxley (2018) find evidence Belgium’s mid-century potato famine impaired adolescent growth using a prison sample from Bruges.

**Figure F1:**
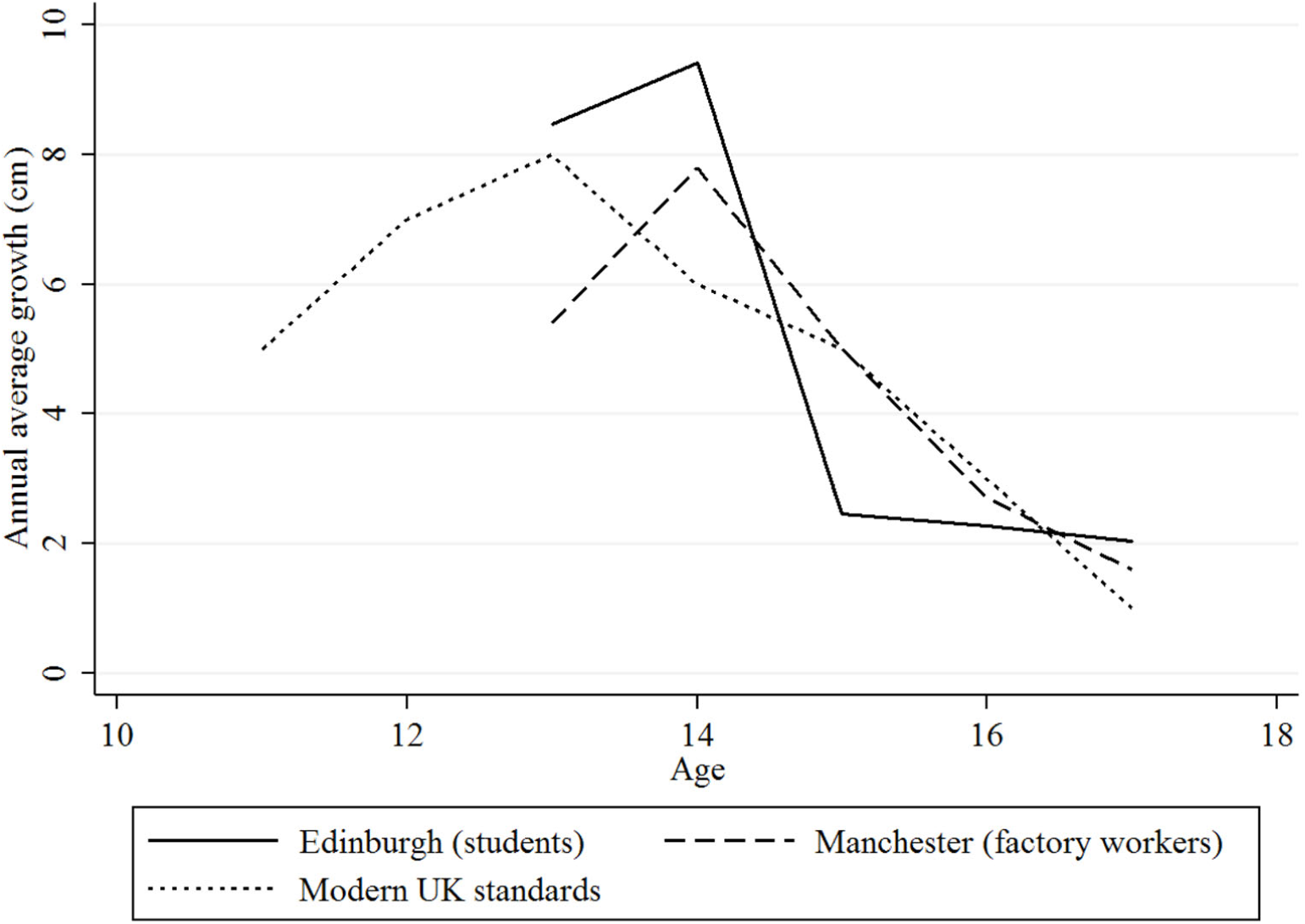
Growth velocity of adolescent boys in the United Kingdom *Sources:* Modern growth charts of boys in the UK are taken from RCPCH (2012); historical growth charts are derived from historical samples of students enrolled in the University of Edinburgh (Forbes 1837), and factory workers from Manchester and Stockport (Quetelet 1842), as discussed in Blum and McLaughlin (2019).

We wish to cross-check our principal findings by ascertaining whether there is a famine shock to adolescent boys, and whether this differed across Ireland. We therefore look at the possibility of growth stunting among those individuals who experienced the Famine during adolescence. We draw new samples of individuals who at the end of the Famine, in 1852, were aged 3-7, 8-12, and 15-17. The first category, aged 3-7, were *born* during the Famine,^29^ while the last, aged 15-17, constitutes the group which was due to undergo its adolescent growth spurt *during* the Famine years.^30^ The middle category serves as a reference group.

Throughout all our tests, reported in Table F1, the only statistically significant famine effect which we can detect among adolescents is found in Dublin-born individuals imprisoned in Dublin. This cohort is approximately 1.0 cm shorter on average than the reference group in our OLS setting (see Section 6 for methodology). However, this result largely disappears when we use our weighted OLS estimator (see Appendix D for methodology). These findings are consistent with our earlier results in that Dublin-born survivors of the Famine show a famine scarring effect, while survivors born in Tipperary do not show any such signs.

**Table F1:** Adolescence famine exposure, OLS and weighted specifications, 5-year famine window

*Panel A: Kilmainham Gaol (males)*
| Measurement: |  |  | Dublin |  |  |  |  |  |  |  |
| --- | --- | --- | --- | --- | --- | --- | --- | --- | --- | --- |
| Sample origin: |  |  | Dublin |  | Leinster |  | Ireland (excl. Dublin) |  | Ireland (excl. Leinster) |  |
| Estimator: |  |  | OLS | Weighted OLS | OLS | Weighted OLS | OLS | Weighted OLS | OLS | Weighted OLS |
|  |  |  | (1) | (2) | (3) | (4) | (5) | (6) | (7) | (8) |
| Constant |  |  | 167.48***<br>(418.15) | 167.33***<br>(368.58) | 169.83***<br>(599.50) | 169.86***<br>(1,053.72) | 168.91***<br>(372.11) | 169.00***<br>(311.41) | 164.56***<br>(292.02) | 163.78***<br>(207.72) |
| *Birth period* | *Age in 1845* | *Age in 1852* |  |  |  |  |  |  |  |  |
| 1835 - 1839 | 6-10 | 13-17 | -1.00***<br>(-3.07) | -0.51<br>(-1.33) | -0.45<br>(-1.24) | -0.24<br>(-1.53) | -0.04<br>(-0.15) | -0.07<br>(-0.32) | -0.54<br>(-0.91) | -0.10<br>(-0.17) |
| 1840 - 1844 | 1-5 | 8-12 | *reference* | *reference* | *reference* | *reference* | *reference* | *reference* | *reference* | *reference* |
| 1845 - 1849<br>(core famine) | unborn | 3-7 | -0.86***<br>(-3.13) | -0.96***<br>(-2.96) | -0.46<br>(-1.64) | -0.67***<br>(-3.36) | 0.18<br>(0.69) | -0.05<br>(-0.19) | 0.32<br>(0.79) | 0.59<br>(1.30) |
| Observations |  |  | 3,165 | 3,165 | 5,275 | 5,275 | 2,974 | 2,974 | 864 | 864 |
| R-squared |  |  | 0.06 | 0.07 | 0.07 | 0.08 | 0.08 | 0.08 | 0.14 | 0.15 |

*Panel B: Clonmel Gaol (males)*
| Measurement: |  |  | Clonmel |  |  |  |  |  |  |  |
| --- | --- | --- | --- | --- | --- | --- | --- | --- | --- | --- |
| Sample origin: |  |  | Tipperary |  | Munster |  | Ireland (excl. Dublin) |  | Ireland (excl. Munster) |  |
| Estimator: |  |  | OLS | Weighted OLS | OLS | Weighted OLS | OLS | Weighted OLS | OLS | Weighted OLS |
|  |  |  | (9) | (10) | (11) | (12) | (13) | (14) | (15) | (16) |
| Constant |  |  | 171.00***<br>(247.45) | 171.03***<br>(228.45) | 169.16***<br>(238.65) | 168.03***<br>(235.72) | 166.57***<br>(217.78) | 166.73***<br>(261.98) | 167.82***<br>(62.24) | 165.36***<br>(101.65) |
| *Birth period* | *Age in 1845* | *Age in 1852* |  |  |  |  |  |  |  |  |
| 1835 - 1839 | 6-10 | 13-17 | -0.22<br>(-0.45) | -0.37<br>(-0.71) | -0.21<br>(-1.22) | -0.32<br>(-1.43) | -0.14<br>(-0.62) | -0.36<br>(-1.47) | 0.62<br>(0.27) | 0.09<br>(0.04) |
| 1840 - 1844 | 1-5 | 8-12 | *reference* | *reference* | *reference* | *reference* | *reference* | *reference* | *reference* | *reference* |
| 1845 - 1849<br>(core famine) | unborn | 3-7 | 0.64<br>(1.46) | 0.36<br>(0.77) | 0.37<br>(1.34) | 0.02<br>(0.07) | 0.31<br>(1.01) | -0.09<br>(-0.24) | 0.80<br>(0.66) | 0.37<br>(0.19) |
| Observations |  |  | 969 | 969 | 1,195 | 1,195 | 1,289 | 1,289 | 149 | 119 |
| R-squared |  |  | 0.02 | 0.02 | 0.03 | 0.03 | 0.06 | 0.07 | 0.26 | 0.32 |

*Notes:* The dependent variable is final adult height. Control variables include age at measurement, time of imprisonment (prison regime dummies), religion, literacy status, birth-county fixed effects and the type of offence. Robust t-statistics in parentheses; where relevant, standard errors are corrected for clustering and autocorrelation by clustering at the birth-county level; \*\*\* p<0.01, \*\* p<0.05, \* p<0.

### Appendix G: International Migration and Stunting

Ireland had a history of negative net migration during the nineteenth century. Of all European countries, Ireland had one of the highest emigration rates per 1,000 population in the latter stages of the nineteenth century (Hatton and Williamson 1994). The most popular migrant destinations were to Great Britain and the United States, in addition to other English-speaking colonies such as British North America, Australia, New Zealand, and to a much lesser extent South Africa. The US was by far the most popular overseas destination. The Irish population in the US was sizeable: over the period 1871-1911 it averaged 34.89 per cent of the population of Ireland.

**Figure G1:**
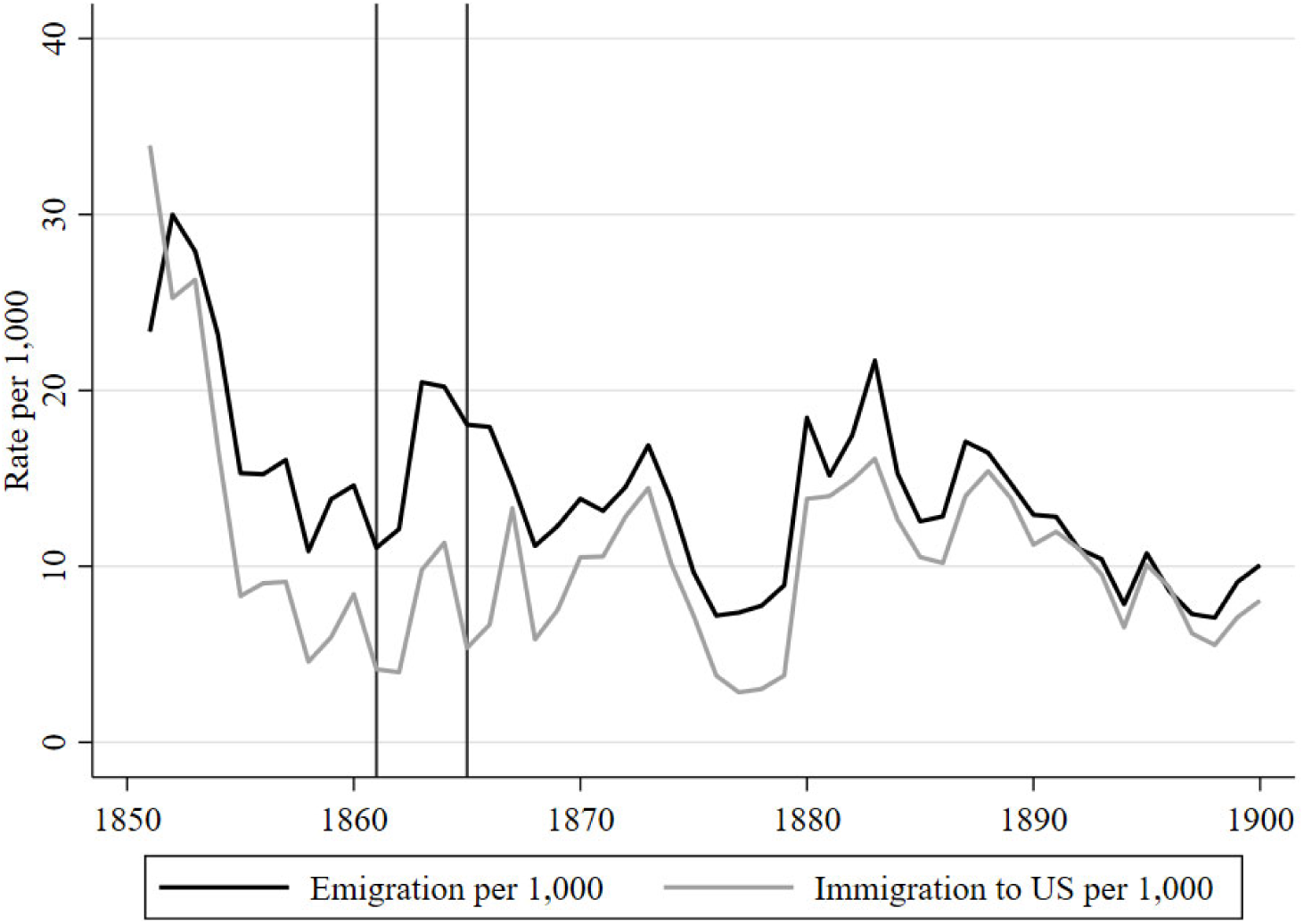
Irish migration rates *Notes*: Immigration refers to Irish immigrants registered in the United States. *Sources*: Department of Social Welfare (1955).

One way to understand the scarring and selection mechanisms in Ireland is to observe the Famine-born cohort overseas. We use data from the Fogel et al. (1990) study of Union Army soldiers to assess a sample of Irish emigrants to the US and compare emigrants’ height with height of Irishmen who remained in Ireland. Here we wade into the Bodenhorn et al. (2017) sample selection debate, although we are supported in the (re-)use of this data source by Komlos and A’Hearn’s (2019) response. We focus on migrant populations within this dataset and compare Irish with British and German migrants.

The Union Army solder database provides data by age and year of enlistment. We focus on birth period between the 1810s and 1840s to investigate whether the Irish-born Union Army soldiers who were born during the Famine show any signs of stunting. Results are presented in Table G1, where male adult height is modelled as a function of birth quinquennium, and age at measurement. We focus on those aged 19 and over to eliminate possible biases resulting from catch-up adolescent growth. We also include two other migrant groups for comparison: soldiers hailing from Great Britain snf the German lands.

Our results suggest that Irish emigrants who were born prior to the famine were taller than their Famine-born counterparts. We use the birth years 1840-1844 as a reference and find that those individuals who were born during the 1845-1849, i.e., those who were experienced famine condition during infancy or early childhood were substantially shorter than the pre-Famine born individuals. The size of this effect is biologically (approximately -2.34 cm) and statistically significant. British migrants born in the period 1845-1849 also experienced some stunting relative to pre-1840s peers, whereas German groups did not.

**Table G1:** Union Army soldiers sample, OLS specifications, 5-year window

| Measurement: | Union Army |  |  |
| --- | --- | --- | --- |
| Sample origin: | Ireland | Great Britain | German lands |
|  | (1) | (2) | (3) |
| Constant | 170.19***<br>(324.10) | 170.02***<br>(205.36) | 168.67***<br>(246.00) |
| *Birth period* |  |  |  |
| 1815 - 1820 | 0.45<br>(0.65) | 0.03<br>(0.03) | 0.92<br>(1.01) |
| 1820 - 1824 | 0.24<br>(0.40) | -0.17<br>(-0.18) | 0.63<br>(0.84) |
| 1825 - 1829 | 1.06*<br>(1.85) | 0.14<br>(0.15) | 1.35*<br>(1.94) |
| 1830 - 1834 | 0.83<br>(1.45) | -0.00<br>(-0.00) | 0.78<br>(1.12) |
| 1835 - 1839 | 0.70<br>(1.47) | 0.53<br>(0.74) | 0.95<br>(1.58) |
| 1840 - 1844 | *reference* | *reference* | *reference* |
| 1845 - 1849 (core famine) | -2.34***<br>(-3.44) | -1.75*<br>(-1.66) | 0.64<br>(0.77) |
| Observations | 2,977 | 1,329 | 2,672 |
| R-squared | 0.03 | 0.03 | 0.01 |

*Notes:* The dependent variable is final adult height. Control variables constitute age at measurement, and year of enlistment. Robust t-statistics in parentheses. \*\*\* p<0.01, \*\* p<0.05, \* p<0.1.

## Government Publications

British Parliamentary Papers [BPP] (1823). *Report of inspectors general on the general state of the prisons of Ireland*. (342).

____ (1826). *Fourth report of the inspectors general on the general state of the prisons of Ireland*. (173).

____ (1840). *Eighteenth report of the inspectors general on the general state of the prisons of Ireland, 1839*. [240].

____ (1843). *Twenty-first report of the inspectors-general on the general state of the prisons of Ireland, 1842*. [462].

____ (1847-48). *Twenty-sixth report of the inspectors-general on the general state of the prisons of Ireland, 1847*. [952].

____ (1849). *Twenty-seventh report of the Inspectors-General on the general state of the prisons of Ireland, 1848*. [1069].

____ (1851). *Abstract of census of Ireland, 1841 and 1851*. (673).

____ (1865). *Judicial statistics, 1864, Ireland*. [3563].

____ (1867). *Forty-fifth report of the inspectors-general on the general state of the prisons of Ireland, 1866*. [3915].

____ (1867-68). *Judicial statistics, 1867, Ireland*. [4071].

____ (1868-69). *First annual report of the registrar-general of marriages, births, and deaths in Ireland*. [4137]

____ (1871). *Criminal and Judicial Statistics, 1871*. [C.674].

____ (1873). *Fifty-first report of the Inspectors-General on the general state of the prisons of Ireland, 1872* .[C. 837].

____ (1873). *Criminal and judicial statistics, 1872, Ireland*. [C.851].

____ (1874). *Fifty-second report of the Inspectors-General on the general state of the prisons of Ireland, 1873*. [C. 966].

____ (1874). *Eighth annual report of the Registrar-General of the marriages, births, and deaths in Ireland*. [C.968].

____ (1874). *Criminal and judicial statistics, 1873, Ireland*. [C.1034].

____ (1875). *Fifty-third report of the inspectors-general on the general state of the prisons of Ireland, 1874*. [C.1256].

____ (1877). *Drunkenness (Ireland)*. (124).

____ (1878-79). *First report of the General Prisons Board, Ireland, 1879; with appendices*. [C.2447]

____ (1882). *Criminal and judicial statistics, 1881, Ireland*. [C.3355].

____ (1884). *Summary proceedings for drunkenness in 1883 (Ireland)*. (1).

____ (1888). *Copy of Rules and Regulations in Force in the Prisons in Ireland*. (329).

____ (1889). *Return of the Cost of Police*. (350).

____ (1890). *Twelfth report of the General Prisons Board, Ireland, 1889-90*. [C.6182].

____ (1892). *Criminal and judicial statistics, Ireland, 1891*. [C.6782].

____ (1900). *Twenty-second report of the General Prisons Board, Ireland, 1899-1900*. [Cd.293].

____ (1902). *Judicial statistics, Ireland, 1901*. [Cd. 1208, 1187].

____ Department of Social Welfare [Republic of Ireland] (1955). *Report of Commission on Emigration and Other Population Problems*. pp. 1948–1954 R. 84 (Pr. 2541).

## Footnotes

1 A literature suggests height proxies related traits: cognitive abilities (Schick and Steckel 2015), mental health (Case and Paxson 2008a), and the ability to generate a wage premium (Persico et al. 2004; Paxson et al. 2009).

2 This depends on the overall disease environment, the mother’s diet, the placenta’s ability to supply nutrients, and even the mother’s own *in utero* experience (Calkins and Devaskar 2011). Prenatal shocks can be counteracted by compensatory catch-up growth, but this is still associated with negative adult health outcomes (Barker 2004: p. 592S).

3 We find a similar pattern among those who experienced this famine during adolescence: scarring in the urban-born sample, but selection among rural-born individuals. And the permanent change in nutritional environment brought on by the Famine resulted in a further decline of 0.7 to 1.2 cm for the post-Famine generation

4 Kim et al. (2013) focus on second-generation human capital outcomes from the Great Leap Forward famine, using data from the 2000 census. Their identification strategy instruments for the estimated death rates of place of parental birthplace and they find significant second-generation effects in terms of secondary school attendance.

5 These alternative estimators include: the deviation from cohort trend which considers outlying cohorts; a difference-in-difference estimator which exploits regional variation in famine severity; and an instrumental variable which uses official exaggerations of grain yields as instruments for famine severity.

6 Forced migration to Australia (i.e., the transportation of convicts) was no longer practiced in the period under investigation.

7 970 skeletal remains were exhumed and examined, representing just under half of the known deaths from the incomplete workhouse records (estimated 2,234 deaths) (Geber 2016).

8 33% of the skeletal remains were aged under five and 48% were under 12 (Geber 2011).

9 Beaumont et al. (2013) compare the Kilkenny sample with a sample from the Lukin Street Catholic graveyard in London. They find the Kilkenny sample had a lower d15n value, indicating lower variance in diet. However, the lack of supporting epigraphic evidence renders it difficult to establish whether the skeletal remains were native-born Londoners, first or second-generation Irish, or local-born spouses of Irish (cf. Morgan 2013).

10 For Clonmel the years of incarceration are 1840 to 1928, and for Kilmainham it is 1798 to 1910. We exploit only those records pertaining to individuals incarcerated after the Famine had ended.

11 We follow Ó Gráda’s (1991, 1994, 1996) use of prison registers from Dublin’s Kilmainham Gaol and Tipperary’s Clonmel Gaol. While Ó Gráda’s studies examine criminals *imprisoned* during the Famine, we look at those *born* during the Famine and institutionalised later.

12 Drunkards represent approximately 40% of our sample, while 41% of the all prisoners in Ireland were drunkards in 1878-79 (BPP 1878-79, pp. 40-41), and 47% in 1889-90 (BPP 1890, pp. 38-39). See Appendix A for further discussion.

13 The cumulative figure for transportation is eye-catching, but it was less dramatic on an annual basis; over the period 1839-44 transportation accounted for 9% of all sentences.

14 All prisoners’ heights were recorded in Imperial measurements using a yardstick, a vertically-mounted measuring rod. We convert all measurements to the metric system to aid with interpretation.

15 For exemplary studies on this widely-observed phenomenon, see Martínez-Carrión and Moreno-Lázaro (2007) on Spain; Riggs and Cuff (2013) on Scotland; Baten (2009) on Bavaria; and Zehetmayer (2017) on the US.

16 Literacy levels were relatively high given the existence of state-funded education dating from the 1830s (Blum et al. 2017).

17 We categorise recidivists by the first crime for which they were imprisoned.

18 Similarly, accounting for the effect of shrinking among individuals aged 50 and above – estimated by Fernihough and McGovern (2015) at 0.09% per year – allows us to extend average height trends further back in time.

19 For a full exposition of the quantile regression methodology, see Buchinsky (1994).

20 We report only key indicators, such as the estimated height change due to the Famine.

21 Approximately 80% of an individual’s height is determined genetically (Silventoinen 2003). We therefore expect the R-squared value of 0.2 is the maximum theoretically possible obtainable value for this goodness-of-fit estimator.

22 Indeed, mean height for adults imprisoned in Kilmainham is lower than mean height in Clonmel (Table 4).

23 The only battlegrounds of Glorious Revolution were in Ireland.

24 Catholics faced discrimination under various Penal Laws, although their enforcement varied. For example, Catholics had restrictions placed on the ownership and inheritance of land, were barred from holding most public offices, and did not have the right to sit in Parliament until the passing of the Catholic Emancipation Act in 1829.

25 Case and Paxson (2008b) find individual height reflects superior intellectual capabilities, and argue the latter explain a large portion of the height premium paid in labour markets.

26 McCrea (2016) conducts a geographic analysis of the prisoners in our dataset hailing from Dublin at the time of incarceration. She finds they overwhelmingly came from poorer parts of the city, such as inner-city slums with poor public health provisioning. She also finds this did not change much across time: Kilmainham’s inmates likely remained poor. It is exactly these individuals who we would expect to have been most affected by famine conditions.

27 Precise dating of the Famine is complicated as it has no clear beginning or end date (Ó Gráda 1999, pp. 37-38). If a famine is defined as excess mortality, then the start date could not be before the autumn of 1846, although contemporaries were aware of harvest shortfalls in the autumn of 1845. Dating the end date is equally problematic – compare, e.g., Woodham-Smith (1991) with Edwards and O’Neill (1956) – with end-dates of studies ranging from 1849

28 In common with the regression analysis in the rest of this paper, we control for various personal characteristics,

29 Of the 3,165 individuals born and incarcerated in Dublin, 37.1% belong to this 3-7 category. And for Tipperary-born individuals incarcerated in Clonmel, this is 51.7%.

30 27.0% of Dublin-born Dublin prisoners belong to this 15-17 category, and 19.6% of Tipperary-born Clonmel prisoners.

